# Development and Validation of a Simplified Risk Score for the Prediction of Critical COVID-19 Illness in Newly Diagnosed Patients

**DOI:** 10.1101/2021.02.07.21251260

**Authors:** Stanislas Werfel, Carolin E. M. Jakob, Stefan Borgmann, Jochen Schneider, Christoph Spinner, Maximilian Schons, Martin Hower, Kai Wille, Martina Haselberger, Hanno Heuzeroth, Maria M. Rüthrich, Sebastian Dolff, Johanna Kessel, Uwe Heemann, Jörg Janne Vehreschild, Siegbert Rieg, Christoph Schmaderer, on behalf of the LEOSS study group

## Abstract

Scores for identifying patients at high risk of progression of the coronavirus disease 2019 (COVID-19), caused by the Severe Acute Respiratory Syndrome Coronavirus 2 (SARS-CoV-2), are discussed as key instruments for clinical decision-making and patient management during the current pandemic.

Here we used the patient data from the multicenter Lean European Open Survey on SARS-CoV-2 - Infected Patients (LEOSS) and applied a technique of variable selection in order to develop a simplified score to identify patients at increased risk of critical illness or death.

A total of 1,946 patients, who were tested positive for SARS-CoV-2 were included in the initial analysis. They were split into a derivation and a validation cohort (n=1,297 and 649, respectively). A stability selection among a total of 105 baseline predictors for the combined endpoint of progression to critical phase or COVID-19-related death allowed us to develop a simplified score consisting of five predictors: CRP, Age, clinical disease phase (uncomplicated vs. complicated), serum urea and D-dimer (abbreviated as CAPS-D score). This score showed an AUC of 0.81 (CI95%: 0.77-0.85) in the validation cohort for predicting the combined endpoint within 7 days of diagnosis and 0.81 (CI95%: 0.77-0.85) during the full follow-up. Finally, we used an additional prospective cohort of 682 patients, who were diagnosed largely after the “first wave” of the pandemic to validate predictive accuracy of the score, observing similar results (AUC for an event within 7 days: 0.83, CI95%, 0.78-0.87; for full follow-up: 0.82, CI95%, 0.78-0.86).

We thus successfully establish and validate an easily applicable score to calculate the risk of disease progression of COVID-19 to critical illness or death.

## Introduction

The first human cases of Coronavirus disease 2019 (COVID-19) were described in December 2019 in Wuhan^1^. From that on, COVID-19 has developed to one of the most disastrous pandemics that we have experienced in our civilization since the Spanish flu in the beginning of 20^th^ century^2,3^. Exponential spread of the disease-causing Severe Acute Respiratory Syndrome Coronavirus 2 (SARS-CoV-2), as has happened throughout Europe during the first wave of the pandemic, can result in an excessive hospital overload and a shortage of healthcare resources, which may lead to a negative impact on patient outcomes^4^. This experience underpinned the importance of an effective process to allocate limited health care resources towards COVID-19 patients who most likely benefit from them. Consequently, to guarantee functional patient care, disease severity assessment for patients presenting to the emergency department (ED) may prove very helpful and guide frontline physicians in the decision-making process. On the one hand, a high number of patients deteriorate rapidly after hospital admission and require transfer to the Intensive care unit (ICU), while on the other hand clinical conditions of other COVID-19 patients improve quite fast. In this respect, a prediction model can support physicians at determining if patients require hospital admission or can be can be followed up in outpatient care.

A risk assessment score may additionally prove to be a helpful tool for estimating the individual risk benefit tradeoff for therapeutic interventions.

The aim of the current study was to develop a simplified risk prediction model based on clinical and demographic characteristics and laboratory findings present at the time point of COVID-19 diagnosis to estimate the risk for clinical deterioration to critical illness. To this end we use data from the Lean European Open Survey on SARS-CoV-2 (LEOSS) project – a prospective European multi-center cohort study^5^.

## Methods

### Study design and patient cohort

This analysis includes patients who received care at a LEOSS partner site (as inpatient or outpatient) starting March 16, 2020. Cases documented in the LEOSS registry up to August 6, 2020 contributed to the initial cohort, which was split into a derivation and validation sets. Cases entered from August 7, 2020 to November 18, 2020 contributed to additional test sets (Figure 2A). The design of the LEOSS study and data acquisition have been described previously^5^.

Data were recorded anonymously and no patient-identifying data were stored. Written patient informed consent was waived. Continuous parameters were categorized. In order to ensure anonymity in all steps of the analysis process, an individual LEOSS Scientific Use File (SUF) was created, which is based on the LEOSS Public Use File (PUF) principles, as described previously^5^. Following these principles, for a minor portion of patients and variables values were removed from the dataset and set to missing to ensure anonymization. Approval for LEOSS was obtained by the applicable local ethics committees of all participating centers and the study was registered at the publicly accessible German Clinical Trails Register (DRKS, No. DRKS00021145).

All predictors included in the stability selection are listed in the Table 1 and Supplementary Table 1. We predefined a combined endpoint of progression to critical disease or COVID-19-related death. Definition of the disease phases is summarized in Figure 1. Baseline (day 0) was defined as the day of the first positive SARS-CoV-2 testing. Only baseline predictors were included in the analysis (for lab values collected within 48 hours of diagnosis). If no CT was conducted within 48 hours of positive testing, we made an exception and included those CT-scan variables collected after this time but during the same clinical phase which was present at baseline. For the analysis we additionally calculated a separate predictor describing if the patient has any cardiovascular (CV) comorbidity, defined as any of the following being present: history of (H/O) myocardial infarction, aortic stenosis, atrioventricular (AV) block, carotid artery disease, chronic heart failure, peripheral vascular disease, hypertension, atrial fibrillation (AF) or coronary artery disease. An additional variable was also calculated for any neurologic comorbidity, defined as any of the following being reported for the patient: hemiplegia, dementia, cerebrovascular disease or stroke, multiple sclerosis, myasthenia gravis, neuromyelitis optica spectrum disorder (NMOSD), movement disorder (e.g. Parkinson’s disease, Dystonia, Ataxia, Tremor), motoneuron diseases (e.g. amyotrophic lateral sclerosis, spinal muscular atrophy), other neurological autoimmune diseases, other prior neurological diagnosis. Lastly, we defined a predictor for any malignant neoplastic disease as any of the following being reported: H/O lymphoma, leukemia, solid tumor, solid metastasized tumor, stem cell transplantation.

**Table 1.**
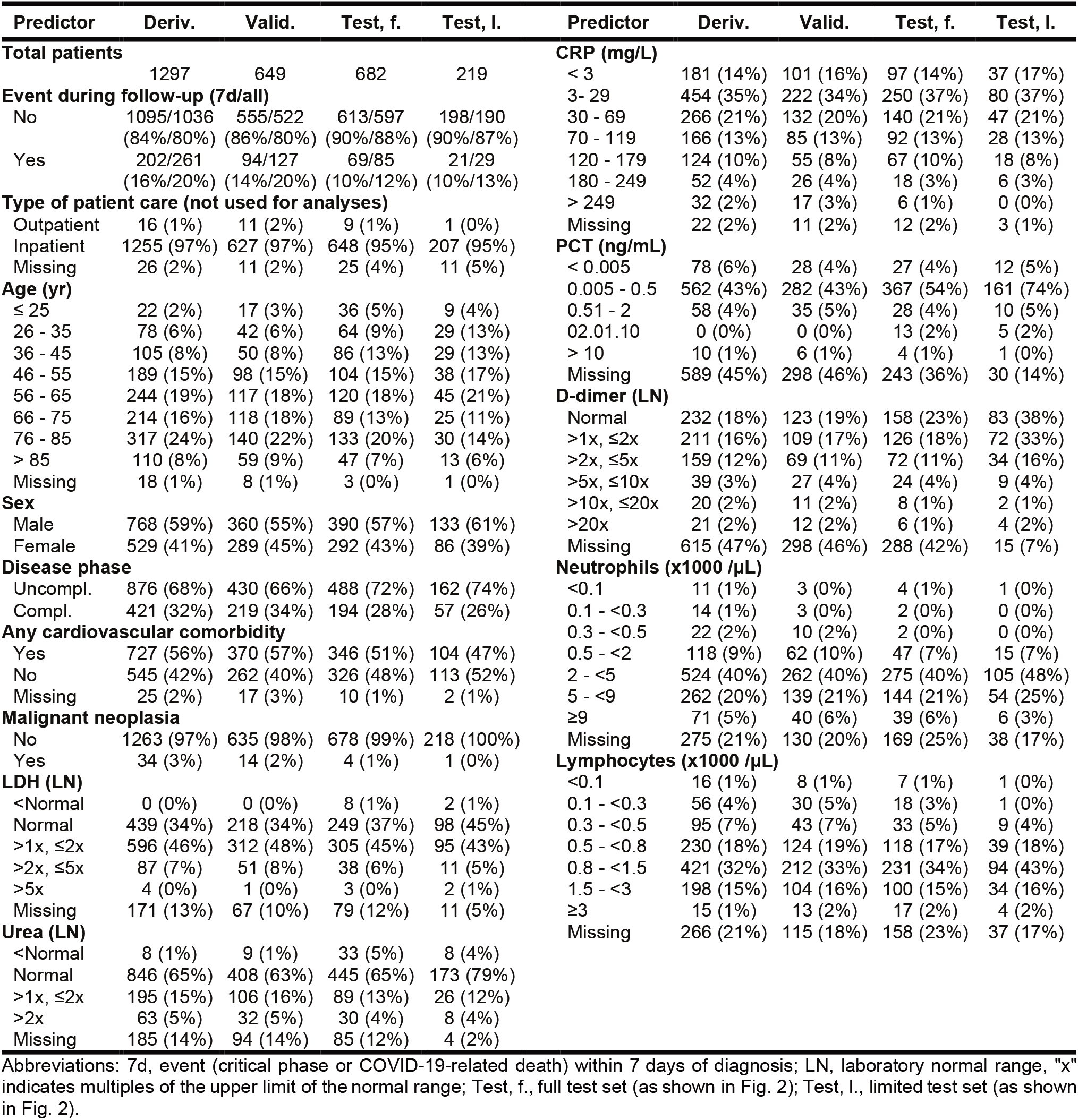
Characteristics of patients in the derivation and validation datasets.

| Predictor | Deriv. | Valid. | Test, f. | Test, l. | Predictor | Deriv. | Valid. | Test, f. | Test, l. |
| --- | --- | --- | --- | --- | --- | --- | --- | --- | --- |
| **Total patients** |  |  |  |  | **CRP (mg/L)** |  |  |  |  |
|  | 1297 | 649 | 682 | 219 | < 3 | 181 (14%) | 101 (16%) | 97 (14%) | 37 (17%) |
| **Event during follow-up (7d/all)** |  |  |  |  | 3- 29 | 454 (35%) | 222 (34%) | 250 (37%) | 80 (37%) |
| No | 1095/1036 | 555/522 | 613/597 | 198/190 | 30 - 69 | 266 (21%) | 132 (20%) | 140 (21%) | 47 (21%) |
|  | (84%/80%) | (86%/80%) | (90%/88%) | (90%/87%) | 70 - 119 | 166 (13%) | 85 (13%) | 92 (13%) | 28 (13%) |
| Yes | 202/261 | 94/127 | 69/85 | 21/29 | 120 - 179 | 124 (10%) | 55 (8%) | 67 (10%) | 18 (8%) |
|  | (16%/20%) | (14%/20%) | (10%/12%) | (10%/13%) | 180 - 249 | 52 (4%) | 26 (4%) | 18 (3%) | 6 (3%) |
| **Type of patient care (not used for analyses)** |  |  |  |  | > 249 | 32 (2%) | 17 (3%) | 6 (1%) | 0 (0%) |
| Outpatient | 16 (1%) | 11 (2%) | 9 (1%) | 1 (0%) | Missing | 22 (2%) | 11 (2%) | 12 (2%) | 3 (1%) |
| Inpatient | 1255 (97%) | 627 (97%) | 648 (95%) | 207 (95%) | **PCT (ng/mL)** |  |  |  |  |
| Missing | 26 (2%) | 11 (2%) | 25 (4%) | 11 (5%) | < 0.005 | 78 (6%) | 28 (4%) | 27 (4%) | 12 (5%) |
| **Age (yr)** |  |  |  |  | 0.005 - 0.5 | 562 (43%) | 282 (43%) | 367 (54%) | 161 (74%) |
| ≤ 25 | 22 (2%) | 17 (3%) | 36 (5%) | 9 (4%) | 0.51 - 2 | 58 (4%) | 35 (5%) | 28 (4%) | 10 (5%) |
| 26 - 35 | 78 (6%) | 42 (6%) | 64 (9%) | 29 (13%) | 02.01.10 | 0 (0%) | 0 (0%) | 13 (2%) | 5 (2%) |
| 36 - 45 | 105 (8%) | 50 (8%) | 86 (13%) | 29 (13%) | > 10 | 10 (1%) | 6 (1%) | 4 (1%) | 1 (0%) |
| 46 - 55 | 189 (15%) | 98 (15%) | 104 (15%) | 38 (17%) | Missing | 589 (45%) | 298 (46%) | 243 (36%) | 30 (14%) |
| 56 - 65 | 244 (19%) | 117 (18%) | 120 (18%) | 45 (21%) | **D-dimer (LN)** |  |  |  |  |
| 66 - 75 | 214 (16%) | 118 (18%) | 89 (13%) | 25 (11%) | Normal | 232 (18%) | 123 (19%) | 158 (23%) | 83 (38%) |
| 76 - 85 | 317 (24%) | 140 (22%) | 133 (20%) | 30 (14%) | >1x, ≤2x | 211 (16%) | 109 (17%) | 126 (18%) | 72 (33%) |
| > 85 | 110 (8%) | 59 (9%) | 47 (7%) | 13 (6%) | >2x, ≤5x | 159 (12%) | 69 (11%) | 72 (11%) | 34 (16%) |
| Missing | 18 (1%) | 8 (1%) | 3 (0%) | 1 (0%) | >5x, ≤10x | 39 (3%) | 27 (4%) | 24 (4%) | 9 (4%) |
| **Sex** |  |  |  |  | >10x, ≤20x | 20 (2%) | 11 (2%) | 8 (1%) | 2 (1%) |
| Male | 768 (59%) | 360 (55%) | 390 (57%) | 133 (61%) | >20x | 21 (2%) | 12 (2%) | 6 (1%) | 4 (2%) |
| Female | 529 (41%) | 289 (45%) | 292 (43%) | 86 (39%) | Missing | 615 (47%) | 298 (46%) | 288 (42%) | 15 (7%) |
| **Disease phase** |  |  |  |  | **Neutrophils (x1000 /μL)** |  |  |  |  |
| Uncompl. | 876 (68%) | 430 (66%) | 488 (72%) | 162 (74%) | <0.1 | 11 (1%) | 3 (0%) | 4 (1%) | 1 (0%) |
| Compl. | 421 (32%) | 219 (34%) | 194 (28%) | 57 (26%) | 0.1 - <0.3 | 14 (1%) | 3 (0%) | 2 (0%) | 0 (0%) |
| **Any cardiovascular comorbidity** |  |  |  |  | 0.3 - <0.5 | 22 (2%) | 10 (2%) | 2 (0%) | 0 (0%) |
| Yes | 727 (56%) | 370 (57%) | 346 (51%) | 104 (47%) | 0.5 - <2 | 118 (9%) | 62 (10%) | 47 (7%) | 15 (7%) |
| No | 545 (42%) | 262 (40%) | 326 (48%) | 113 (52%) | 2 - <5 | 524 (40%) | 262 (40%) | 275 (40%) | 105 (48%) |
| Missing | 25 (2%) | 17 (3%) | 10 (1%) | 2 (1%) | 5 - <9 | 262 (20%) | 139 (21%) | 144 (21%) | 54 (25%) |
| **Malignant neoplasia** |  |  |  |  | ≥9 | 71 (5%) | 40 (6%) | 39 (6%) | 6 (3%) |
| No | 1263 (97%) | 635 (98%) | 678 (99%) | 218 (100%) | Missing | 275 (21%) | 130 (20%) | 169 (25%) | 38 (17%) |
| Yes | 34 (3%) | 14 (2%) | 4 (1%) | 1 (0%) | **Lymphocytes (x1000 /μL)** |  |  |  |  |
| **LDH (LN)** |  |  |  |  | <0.1 | 16 (1%) | 8 (1%) | 7 (1%) | 1 (0%) |
| <Normal | 0 (0%) | 0 (0%) | 8 (1%) | 2 (1%) | 0.1 - <0.3 | 56 (4%) | 30 (5%) | 18 (3%) | 1 (0%) |
| Normal | 439 (34%) | 218 (34%) | 249 (37%) | 98 (45%) | 0.3 - <0.5 | 95 (7%) | 43 (7%) | 33 (5%) | 9 (4%) |
| >1x, ≤2x | 596 (46%) | 312 (48%) | 305 (45%) | 95 (43%) | 0.5 - <0.8 | 230 (18%) | 124 (19%) | 118 (17%) | 39 (18%) |
| >2x, ≤5x | 87 (7%) | 51 (8%) | 38 (6%) | 11 (5%) | 0.8 - <1.5 | 421 (32%) | 212 (33%) | 231 (34%) | 94 (43%) |
| >5x | 4 (0%) | 1 (0%) | 3 (0%) | 2 (1%) | 1.5 - <3 | 198 (15%) | 104 (16%) | 100 (15%) | 34 (16%) |
| Missing | 171 (13%) | 67 (10%) | 79 (12%) | 11 (5%) | ≥3 | 15 (1%) | 13 (2%) | 17 (2%) | 4 (2%) |
| **Urea (LN)** |  |  |  |  | Missing | 266 (21%) | 115 (18%) | 158 (23%) | 37 (17%) |
| <Normal | 8 (1%) | 9 (1%) | 33 (5%) | 8 (4%) |  |  |  |  |  |
| Normal | 846 (65%) | 408 (63%) | 445 (65%) | 173 (79%) |  |  |  |  |  |
| >1x, ≤2x | 195 (15%) | 106 (16%) | 89 (13%) | 26 (12%) |  |  |  |  |  |
| >2x | 63 (5%) | 32 (5%) | 30 (4%) | 8 (4%) |  |  |  |  |  |
| Missing | 185 (14%) | 94 (14%) | 85 (12%) | 4 (2%) |  |  |  |  |  |

Abbreviations: 7d, event (critical phase or COVID-19-related death) within 7 days of diagnosis; LN, laboratory normal range, "x" indicates multiples of the upper limit of the normal range; Test, f., full test set (as shown in Fig. 2); Test, l., limited test set (as shown in Fig. 2).

**Figure 1.**
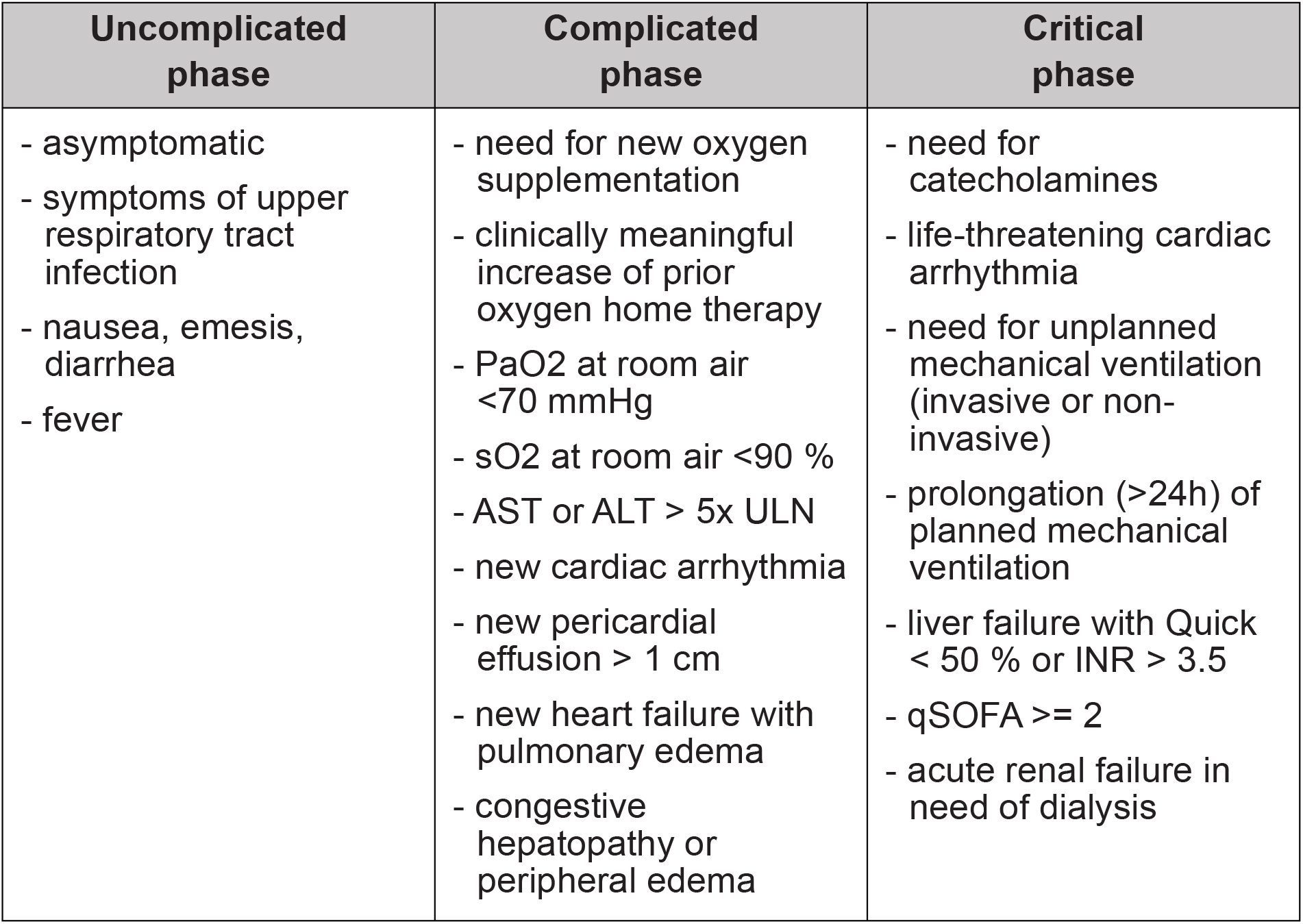
Definition of COVID-19 disease phases in the LEOSS registry. Patients were assigned to the highest phase for which at least one characteristic was fulfilled. Abbreviations: ALT, alanine transaminase; AST, aspartate transaminase; INR, international normalized ratio of prothrombin time; PaO2, partial pressure of oxygen in arterial blood; qSOFA, quick sequential organ failure assessment score; sO2, blood oxygen saturation; ULN, upper limit of normal;

**Figure 2.**
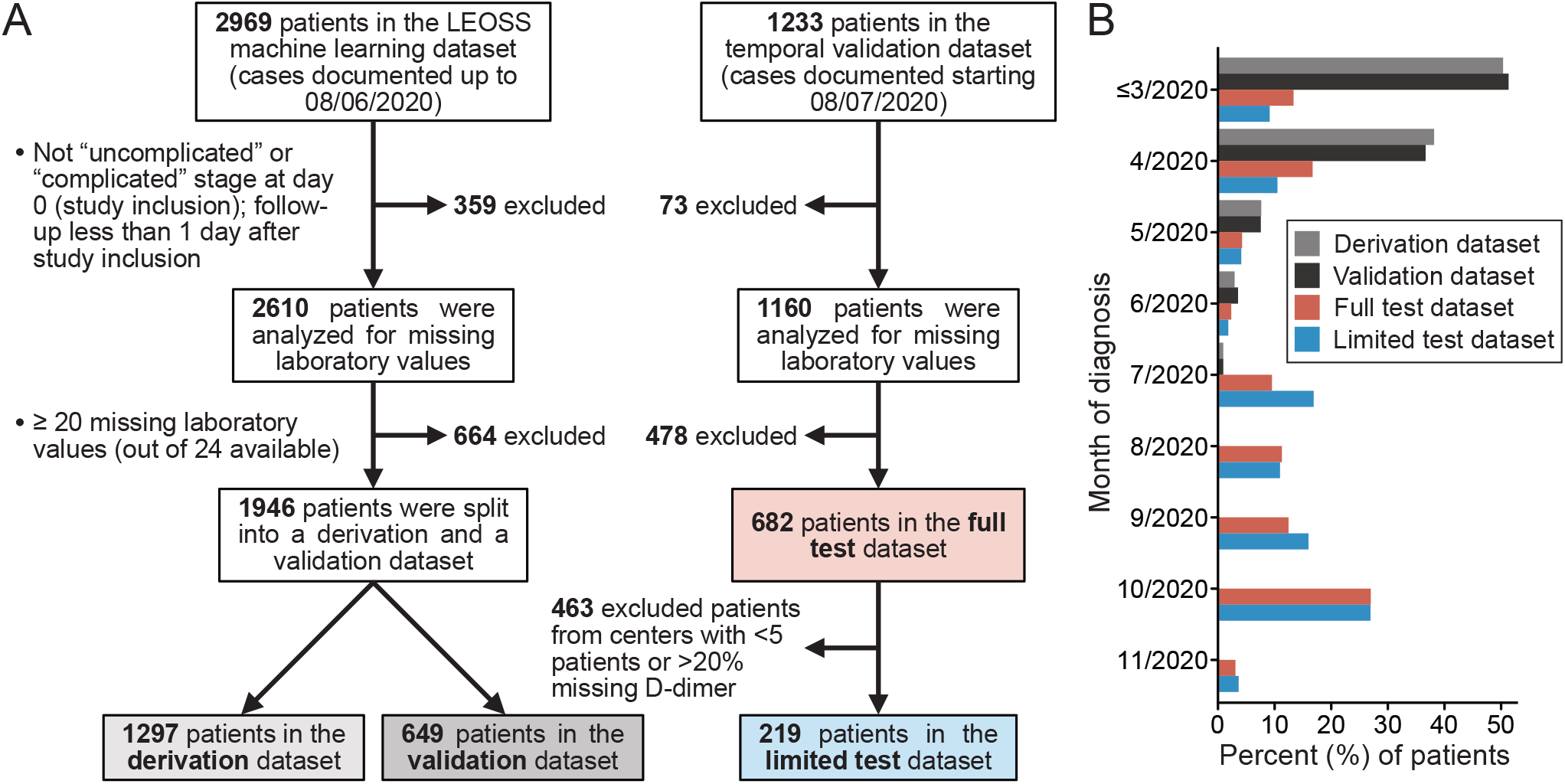
Patient flow diagram (A) and months of COVID-19 diagnosis (B) for the different datasets.

### Statistical analysis

All analyses were carried out in R (version 3.6.3). Random forest analyses (including missing value imputations and individual Boruta stability selection steps) were calculated using the “randomForestSRC” package by Ishwaran et Kogalur^6^.

Among the available baseline variables of the LEOSS dataset (≈170 predictors) we selected those with less than 50% missing values among the combined derivation and validation dataset (n=1946 patients, as shown in Figure 2), with an exception made for Troponin T (52% missing) and pancreas lipase (56% missing). This resulted in a total of 105 predictors (as listed in the Table 1 and Supplementary Table 1). Since the time to event data in the anonymized LEOSS cohort was grouped for patients experiencing an event at ≥8 days after study inclusion, the time variable was coded accordingly as 1 to 7 days and ≥8 days, resulting in 8 bins for the time variable (Supplementary Table 1). These were used for the time-to-event approaches: random survival forest and Cox models and for C-index calculation. Continuous predictors are binned as value ranges in the LEOSS cohort due to anonymization and for the analysis the ranges were coded as consecutively increasing integers.

We performed unsupervised random forest missing value imputation using either the data of the combined derivation and validation datasets (n=1946 patients) or, separately, the full test set (n=682 patients, Figure 2), while withholding the outcome variables. We thus generated 20 imputed datasets for each of the cohorts.

We performed a split into a derivation and a validation cohort with similar characteristics based on the following predefined potential confounders: age, sex, presence of dyspnoe, neutrophil count, lymphocyte count, lactate dehydrogenase (LDH), bilirubin, CRP, PCT, D-dimer, H/O malignant neoplasia, presence of any CV comorbidity (as defined above) and the number of events. To this end we performed 1000 random splits at a 2/3 and 1/3 ratio and calculated for each split and variable the standardized mean difference, selecting the split with the smallest maximal standardized mean difference between these predictors.

Variable selection was carried out using the Boruta algorithm^7^ at 100 iterations using equal proportions of the 20 imputed derivation datasets and a p-value of 0.01 for selection. For classification random forests we used the presence of an event (critical phase or COVID-19-related death) within 7 days of diagnosis as the outcome of interest during Boruta selection. We used the balanced method by Chen at al.^8^ both during Boruta selection and modelling with the selected variables. Likewise, we used survival random forest as described by Ishwaran et al.^9^ both during Boruta selection and during final modelling of time to event data. For survival random forests, since they take time to event into account, also events occurring longer than 7 days after diagnosis were included. Variable importance was calculated using permutation. For Cox and logistic (binomial) regression models we performed ridge (L2) penalization optimized using 20x fold cross-validation on the imputed derivation datasets. Score values were calculated from the ridge penalized binomial regression coefficients of the model containing the five selected predictors on the derivation dataset with missing values replaced with the most common value of the 20 imputed datasets for this patient and predictor and event within 7 days as outcome. Finally, the regression coefficients were divided by the smallest one and rounded to the next whole integer. Two-sided p-values for binomial ridge penalized coefficients were obtained as suggested by Cule et al.^10^, by repeating the ridge regression procedure on a dataset with randomly permuted outcomes 1000 times (using equal amounts of the 20 imputed datasets).

Area under the receiver operating characteristics curve (AUC) and (Harrell’s) C-indices were calculated using linear predictors from the binomial and Cox ridge-penalized regression models or out-of-bag (OOB) predictor estimates for the random forest approaches. 95% confidence intervals for AUC and C-indices were calculated using 1000 bootstraps of patients’ scores using equal contributions of the imputed datasets.

## Results

### Patient population

Important characteristics of the LEOSS cohort were described previously^5^. More diagnosed SARS-CoV-2 cases were available for the current analysis compared to this previous report (2,969 in the first dataset, patients from the first wave of the pandemic, and 1,233 patients in a second test set, Figure 2)^5^. Based on the predefined disease phase (Figure 1) and the availability of laboratory values, a total of 1,946 patients were included in the first round of analysis and split into a derivation and validation groups with similar characteristics (Figure 2). Important characteristics are summarized in Table 1 with a summary of the remaining predictors provided in Supplementary Table 1.

The age distribution in the first dataset was centered, with about equal contribution of patients of ≤65 and >65 years. There were more men than women at 55-59% to 41-45%. At least 56% presented with a known CV comorbidity. The incidence of the combined endpoint, critical phase or COVID-19-related death, within 7 days was 14-16%, and 20% when including any time point during the follow-up (Table 1).

From the second test set (patients whose cases were entered into the registry after the first data export for score derivation) 682 patients passed selection criteria. This set largely consisted of patients diagnosed after June 2020 (Figure 2). Compared to the derivation/validation cohorts, the patients were younger (60% with an age of ≤65 years) and more were diagnosed in an uncomplicated phase (72% vs. 64-68%). Consequently, the event rate was lower with only 10% experiencing an event within 7 days of diagnosis and 12% during the full follow-up (Table 1). Both the derivation and validation datasets consisted almost exclusively of patients receiving inpatient care.

### Predictor selection

We performed Boruta variable stability selection using random forest for classification (RF), resulting in the selection of 5 (out of 105) predictors (Table 2). These were: CRP, disease phase, age, serum urea, and D-dimer (Supplementary Figure 1A). Interestingly, including only these five predictors in a logistic regression model achieved results almost on par with the full set of variables (Table2, “RF Boruta”, Binomial ridge, median AUC=0.81 in the validation cohort).

**Table 2.** Summary of the predictive performances of the analyzed models.

| Selection | Model | N pr. | AUC, 7 d (imp. range) |  | AUC, all (imp. range) |  |
| --- | --- | --- | --- | --- | --- | --- |
|  |  |  | Derivation | Validation | Derivation | Validation |
| All pr. | RF | 105 | 0.83 (0.82-0.83) | 0.83 (0.82-0.83) | 0.83 (0.83-0.83) | 0.83 (0.82-0.83) |
|  | Binomial ridge | 105 | 0.88 (0.86-0.89) | 0.81 (0.80-0.81) | 0.86 (0.86-0.87) | 0.81 (0.80-0.82) |
| RF Boruta | RF | 5 | 0.74 (0.72-0.75) | 0.73 (0.71-0.75) | 0.73 (0.72-0.75) | 0.75 (0.73-0.76) |
|  | Binomial ridge | 5 | 0.80 (0.80-0.80) | 0.81 (0.80-0.81) | 0.80 (0.80-0.80) | 0.81 (0.81-0.82) |
|  | **Score** | **5** | **0.80 (0.80-0.80)** | **0.81 (0.81-0.81)** | **0.80 (0.80-0.80)** | **0.81 (0.81-0.81)** |
|  |  |  | **95%CI, 0.77-0.83** | **95%CI, 0.77-0.85** | **95%CI, 0.77-0.83** | **95%CI, 0.77-0.85** |
|  | Validation on full test set |  |  | **0.83 (0.82-0.83)** |  | **0.82 (0.82-0.82)** |
|  |  |  |  | **95%CI, 0.78-0.87** |  | **95%CI, 0.78-0.86** |
|  | Validation on limited test set |  |  | **0.82 (0.82-0.82)** |  | **0.83 (0.83-0.83)** |
|  |  |  |  | **95%CI, 0.73-0.90** |  | **95%CI, 0.76-0.90** |

Indicated are the median values and the full range for the imputed datasets (in brackets). AUC values were calculated for an event within 7 days of diagnosis ("7 d") and for all time points ("all"). 95% confidence intervals (95%CI) were calculated for score predictions using bootstrapping with equal contributions of the imputed datasets.
Abbreviations: AUC, area under the receiver operating characteristic (ROC) curve; imp., imputation; N pr., number of predictors in the model; pr., predictors; RF, random forest for classification.

We additionally performed Boruta stability selection using a survival random forest (RSF) approach. Here, 24 predictors were retained, with the five predictors from RF Boruta being among the variables with the highest importance (Supplementary Figure 1B). Increasing the number of predictors to 24 had only a minor impact on model performance in the validation dataset compared to five predictors, as measured by Harrell’s C-index (median C-index of 0.77 vs. 0.76 for five predictors, Supplementary Table 2).

### Derivation and validation of a simplified predictive score

Based on the encouraging results and the simple interpretability, we used the coefficients obtained in the binomial ridge regression model with five predictors (Table 3) to derive an additive score for prediction of COVID-19 progression to critical phase or death. The score is outlined in Table 4. It showed a similar performance when compared to the binomial model both in the derivation and the validation datasets (median AUC in the validation dataset for events within 7 days of diagnosis: 0.81, 95% confidence interval (95% CI), 0.77-0.85, and for all events, 0.81, 95% CI, 0.77-0.85, Table 2). Interestingly, the simplified score also showed a similar performance to a Cox regression or an RSF approach with both the five and 24 predictors as measured by Harrell’s C-index (median C-index of 0.76, 95% CI, 0.73-0.80 in the validation cohort, Supplementary Table 2).

**Table 3.** Results of the ridge-penalized binomial regression on the five variables selected by RF Boruta.

| Predictor | Ridge $\beta$ | P-value | Weight |
| --- | --- | --- | --- |
| Age | 0.09 | 0.016 | 1 |
| Disease phase | 0.46 | 0.001 | 5 |
| Urea | 0.30 | 0.010 | 3 |
| CRP | 0.16 | <0.001 | 2 |
| D-dimer | 0.11 | 0.030 | 1 |

**Table 4.**
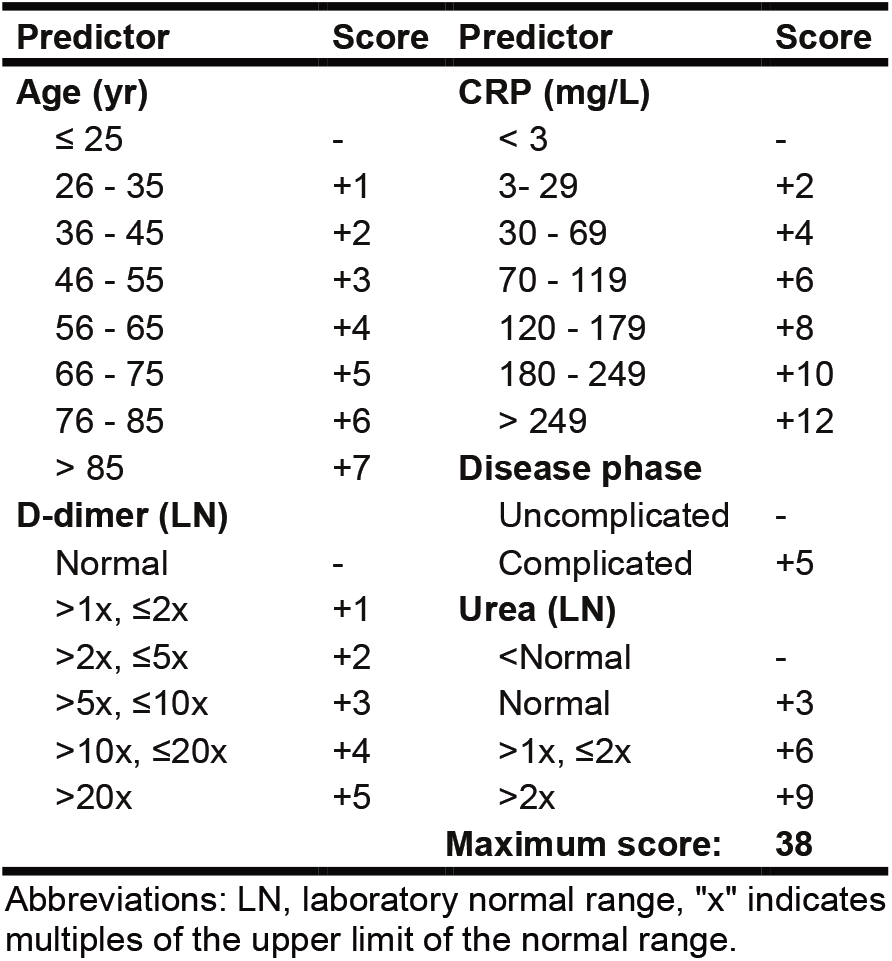
Calculation of the CAPS-D score.

| Predictor | Score | Predictor | Score |
| --- | --- | --- | --- |
| **Age (yr)** |  | **CRP (mg/L)** |  |
| ≤ 25 | - | < 3 | - |
| 26 - 35 | +1 | 3- 29 | +2 |
| 36 - 45 | +2 | 30 - 69 | +4 |
| 46 - 55 | +3 | 70 - 119 | +6 |
| 56 - 65 | +4 | 120 - 179 | +8 |
| 66 - 75 | +5 | 180 - 249 | +10 |
| 76 - 85 | +6 | > 249 | +12 |
| > 85 | +7 | **Disease phase** |  |
| **D-dimer (LN)** |  | Uncomplicated | - |
| Normal | - | Complicated | +5 |
| >1x, ≤2x | +1 | **Urea (LN)** |  |
| >2x, ≤5x | +2 | <Normal | - |
| >5x, ≤10x | +3 | Normal | +3 |
| >10x, ≤20x | +4 | >1x, ≤2x | +6 |
| >20x | +5 | >2x | +9 |
|  |  | **Maximum score:** | **38** |

Abbreviations: LN, laboratory normal range, "x" indicates multiples of the upper limit of the normal range.

As an independent prospective validation group, we used the second test set of patients whose data was entered into the registry after the initial data export (n=682 patients, “full test set” in Figure 2). To further reduce the impact of missing values on the estimation of score performance, we additionally removed patients from centers with >20% missing values for D-dimer, the variable with most missing values (42-47% missing). Centers which enrolled less than five patients were also removed. This resulted in an additional “limited test set” (n=219 patients, Figure 2). This dataset had only a minor portion of missing values (CRP: 1%, serum urea: 2%, D-dimer: 7% missing, Table 2).

In both the full and limited test sets we could confirm a similar performance of the developed score, with a trend towards higher AUC and C-index values compared to the validation dataset (full test set, median AUC for 7 d: 0.83, 95% CI, 0.78-0.87; all events: AUC 0.82, 95% CI, 0.78-0.86; limited test set, median AUC for 7 d: 0.82, 95% CI, 0.73-0.90; all events: AUC 0.83, 95% CI, 0.76-0.90, Table 2; median C-index for full test set: 0.80, 95%CI, 0.76-0.84; limited test set: 0.81, 95%CI, 0.74-0.87, Supplementary Table 2).

Depending on the clinical application, different cutoffs may be considered. We therefore provide the predictive metrics of the score, such as sensitivity, specificity and the positive and negative predictive values (PPV, NPV) vs. the cutoff (Figure 3) as well as the absolute event risks for specific score values (Supplementary Figure 2).

**Figure 3.**
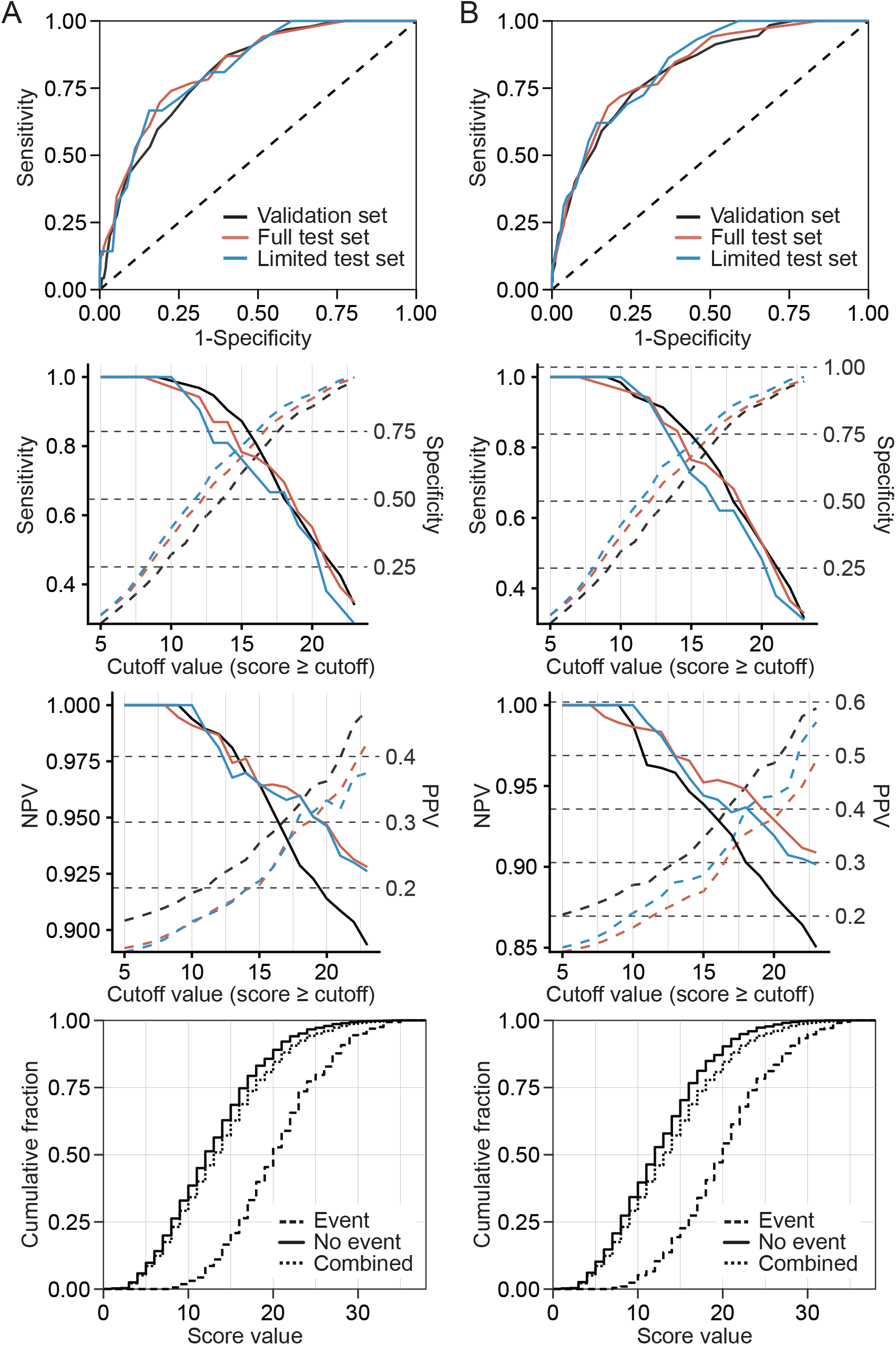
Summary of key characteristics of the score for predicting the combined endpoint of critical phase or COVID-19-related death (A) within 7 days of the diagnosis or (B) at any time point during follow up in the validation and test cohorts. Colour codes distinguish the different datasets as indicated. Sensitivity and NPV are indicted by continuous lines and the corresponding y-axis scaling on the left, while specificity and PPV are indicated by dashed lines and y-axis scaling on the right side of the respective panels. Bottom panels show cumulative fractions of patients meeting respective score cutoffs for a combined validation and full test set (combined n=1331). For all panels the median score (rounded to the next whole integer) of the imputations was calculated for patients with missing values.

Next to the discriminative performance, we observed good calibration with a slope ranging from 0.944 to 1.101 in the different validation/test datasets (Supplementary Figure 3). Interestingly, the Brier score was tendentially smaller in the “full test” compared to the validation dataset (0.076-0.091 vs. 0.106-0.124, Supplementary Figure 3), mirroring the tendency towards a better discriminative performance in this dataset (Table 2 and Supplementary Table 2). Calibration-in-the-large for the “full test” set, which showed a lower event per case rate, was similar to that in the validation set for an event within 7 days (intercept of −0.181 vs. −0.182), with a higher difference for all events (intercept of −0.334 vs. 0.004, potentially reflecting the differences in the event rates between the cohorts).

One method to select a cutoff is by optimizing the modified Youden’s J^11^. For the proposed score the optimal J in the combined validation and full test dataset was at a cutoff of ≥17 both for predictions at 7 days after diagnosis and for all events. Applying this cutoff, on average 69% of patients are predicted to not progress to critical illness (Table 5, combined validation/test dataset) at an NPV of 95% for 7 days after diagnosis and an NPV of 94% for the full follow-up. Patients with scores at or above this threshold had ≈3-fold increased odds of experiencing an event, while patients below this threshold had ≈3-fold decreased odds as measured by the respective likelihood ratios (Table 5).

**Table 5.** Score characteristics at the selected cutoff of ≥17.

|  | **Validation**<br>**set (7d/all)** | **Full test set**<br>**(7d/all)** | **Combined**<br>**(7d/all)** |
| --- | --- | --- | --- |
| Sensitivity | 0.73 / 0.73 | 0.74 / 0.72 | 0.74 / 0.73 |
| Specificity | 0.72 / 0.75 | 0.77 / 0.79 | 0.75 / 0.77 |
| PPV | 0.31 / 0.41 | 0.27 / 0.32 | 0.29 / 0.37 |
| NPV | 0.94 / 0.92 | 0.96 / 0.95 | 0.95 / 0.94 |
| LR+ | 2.6 / 2.9 | 3.3 / 3.3 | 2.9 / 3.1 |
| LR- | 0.37 / 0.36 | 0.34 / 0.36 | 0.35 / 0.36 |
| %score<cutoff | 65% | 72% | 69% |

Abbreviations: %score<cutoff, percentage of patients with scores below the cutoff value ( $\leq 16$ ).; 7d, event (critical disease or COVID-19-related death) within 7 d of diagnosis; all, all events during follow-up; LR+/-, positive/negative likelihood ratio; NPV, negative predictive value; PPV, positive predictive value.

## Discussion

Here we describe the derivation and validation of a COVID-19 risk score for the prediction of the combined end point of critical disease or COVID-19-related death with five predictors. We derive the score in an untargeted way by selecting the most stable predictors among 105 available at baseline in the LEOSS registry in a random forest approach and use regularized regression to calculate the coefficients.

A number of approaches for COVID-19 risk stratification have been reported previously (reviewed by Wynants et al.^12^), several with a similar aim of predicting critical disease, as indicated by admission to the ICU (e.g.^13–15^) or death (e.g. ^14–17^). The availability of factors such as hospital or ICU beds has been limited during the high tide of the pandemic and the resulting strain on health care systems. Thus, difficulties in generalizing outcome predictions obtained under these constraints in currently available scores may arise. In our view some important limiting aspects have to be considered. On the one hand, if hospital beds are limited, the study population for inpatient analyses may overrepresent patients with symptoms of exceptional severity and high risk groups, which may limit generalizability (as e.g. noted for the ISARIC 4C score^16^). Similarly, if ICU resources are limited, the indications for admission may be more conservative, thus a patient may be identified as having a favorable outcome (not being admitted to an ICU) despite having fulfilled clinical criteria at some point. This is exemplified by the finding that one of the predictors included in the COVID-GRAM score was unconsciousness, which may already indicate an outcome of an advanced disease.

Another important consideration is for the generalizability of mortality as an outcome for patient stratification. Case fatality rates have been widely differing across countries^18^, even within the European Union, which may be at least in part attributed to country-specific differences in clinical management of COVID-19 patients and to resource availability during the first wave of the pandemic^4^. This may limit generalizability and potentially require an update to existing scores for mortality prediction^19^ as care providers gain experience with COVID-19 management and the strain on hospitals is reduced.

A previous review on COVID-19 prognosis scores came to an overall negative assessment of the potential bias of these scores and as a result discouraged their use^12^. A combination of characteristics sets apart our approach compared to those available (to our knowledge) at the time of writing and makes it potentially better generalizable for future clinical application: (a) the outcome was not defined in terms of a specific treatment (or lack thereof, i.e. admission to the ICU), but rather based on clinical features (a predefined “critical phase”); (b) the inclusion was based on predefined clinical criteria (“uncomplicated” or “complicated” phase) and (c) the use of a stability selection approach as a means of reducing the number of predictors, as further discussed below. Additionally, the vast majority (>90%) of the cases enrolled into the LEOSS cohort were from Germany^5^, where the capacity of the healthcare system was in general not exceeded during the first wave of the pandemic^20^.

To address bias in predictor selection we used an untargeted approach and resampling techniques (stability selection and cross-validated ridge regression) in order to first internally test the predictions on the derivation dataset and then validate them on a withheld validation cohort. Stability selection helps to ensure the internal validity and sufficient sample size already for the derivation dataset, as a too small sample will reduce variable stability and lead to less variables being selected. Ridge regression shrinks the regression coefficients to achieve improved predictions in a binomial model again with internal (cross-)validation already in the derivation dataset. As a result of the above steps, we could successfully confirm the performance of our score in an independent test set, consisting in its majority of COVID-19 cases diagnosed after the first wave of the pandemic.

An important contributor to the predictive performance of the final score was the predefined clinical phase (“complicated” vs. “uncomplicated”), which summarizes the presence of a manifest organ involvement of the lungs, heart or liver. Of note, some parameters of the complicated phase, such as arterial partial pressure of oxygen (PaO2) and pericardial effusion were acquired by indication (e.g. if an echocardiography or arterial blood gas analysis were performed, but not routinely), therefore for phase assignment these do not have to be taken into account in absence of an indication for the respective measurement.

Serum urea, likely as a measure of kidney involvement, was also an important predictor and outperformed creatinine, as reported previously for mortality^16,21^. This predictor potentially summarizes both preexisting chronic kidney disease (CKD) as a risk factor (as e.g. reported by Williamson et al.^22^ and in Supplementary Figure 1B) and acute kidney injury (AKI) due to COVID-19 as organ involvement (also stable in RSF Boruta, Supplementary Figure 1B). Different mechanisms of AKI in COVID-19 patients have been observed, including indirect involvement e.g. due to a cardiorenal syndrome, direct virus-induced injury as well as immunologic causes such as complement activation (as reviewed in ^23,24^). Differentiating the type of acute kidney involvement in COVID-19 patients may provide further insights and refine risk stratification in future analyses.

Overall, the presented score, despite being limited to only 5 predictors and applying a point system, compared well to more complex prediction models.^14,16^ We suggest a threshold for patients with an increased risk of critical disease at ≥ 17 points, based on the modified Youden’s J. At this threshold we obtained a positive likelihood ratio of 3-fold while retaining a good negative predictive value of 94-95%. Different cutoffs may be considered based on the application and local circumstances (e.g. different local ratio of critical disease per case, travelling time to the next hospital in case of deterioration in an outpatient setting, etc.). The graphs provided in Figure 3 for sensitivity/specificity and PPV/NPV (based on the prevalence in the validation and test datasets) as well as in Supplementary Figure 2 for absolute risk prediction may assist in determining such thresholds.

## Limitations

Our study has limitations. The LEOSS registry is anonymized and continuous parameters were categorized, thus potentially reducing the predictive performance of e.g. laboratory measures. As a real-world dataset, given the heterogeneity of clinical procedures across centers, our analysis had to compensate for missing values. This typically reduces the predictive performance of the respective variables and the probability that they pass stability selection criteria. Thus, some predictors may have been underestimated or missed.

Our analysis was limited to predicting disease progression with information obtained at the time point of first positive SARS-CoV-2 testing (typically occurring during presentation at the medical facility), without taking into account the dynamics of the predictors. In this regard, the days since the beginning of symptoms (uncomplicated phase) to the diagnosis were included as a variable, however it did not pass stability criteria. Also, there were differences between the validation and the test dataset with the latter having a higher proportion of patients diagnosed in the uncomplicated phase (suggesting earlier diagnosis, possibly due to expanded testing capacities after the first wave). Yet, the score still showed a similar or tendentially even better performance in the test set. This indirect evidence suggests that the application of our score may be valid also at time points after diagnosis (or the initial presentation), such as if the patient’s condition or laboratory values deteriorate, however further studies are needed to assess its suitability in such settings.

No information on patient ethnicity/race was available for current analysis, it may be assumed that the distribution follows that in the German population and represents largely Caucasians, which may limit generalizability. External validation in different patient populations is thus needed, also with regard to socioeconomic factors and local standards of care.

Extensive information on comorbid conditions for the study participants was available. Although some passed the criteria in RSF stability selection, none passed RF stability criteria. Yet, having more predictors (24 vs. 5) did not improve the overall predictive performance. This suggests that the increased risk due to these comorbidities may already be reflected by the remaining five predictors (collinearity), therefore relieving the need for inclusion into the score. However, this may not hold true for less common comorbidities, as the overall prediction improvement will be low for low prevalence predictors, even if they have a strong effect for patients suffering from these comorbidities. Thus, a score based on the total population, as presented here, may underestimate high-risk constellations due to rare comorbidities such as specific cancers, autoimmune diseases/immunosuppressive treatments, etc. To our knowledge this limitation applies to most if not all available COVID-19 prognosis scores which were derived on the total population. Yet, such patients may deteriorate rapidly. It seems important to establish the additional risk for specific conditions on top of the used score in future studies.

## Data Availability

Patient data from the LEOSS registry is subject to the LEOSS Governance, Data Use and Access Policy (policy text available on https://leoss.net).

