## Supplementary Material for "Development and Validation of a Simplified Risk Score for the Prediction of Critical COVID-19 Illness in Newly Diagnosed Patients"

### Supplementary Table 1. Patient characteristics (variables not reported in Table 1).

| Predictor | Deriv. | Valid. | Test, f. | Test, l. |
|---|---|---|---|---|
| **Total patients** | | | | |
| | 1297 | 649 | 682 | 219 |
| **Time to critical phase or loss to follow-up** | | | | |
| 1 | 75 (6%) | 40 (6%) | 127 (19%) | 48 (22%) |
| 2 | 82 (6%) | 31 (5%) | 41 (6%) | 17 (8%) |
| 3 | 89 (7%) | 26 (4%) | 38 (6%) | 16 (7%) |
| 4 | 82 (6%) | 39 (6%) | 34 (5%) | 7 (3%) |
| 5 | 84 (6%) | 39 (6%) | 44 (6%) | 12 (5%) |
| 6 | 84 (6%) | 48 (7%) | 34 (5%) | 13 (6%) |
| 7 | 87 (7%) | 39 (6%) | 42 (6%) | 14 (6%) |
| ≥8 | 714 (55%) | 387 (60%) | 322 (47%) | 92 (42%) |
| **Day of symptom/uncompl. phase start (relative to diagnosis)** | | | | |
| ≤ -8 | 188 (14%) | 91 (14%) | 76 (11%) | 24 (11%) |
| -7 | 68 (5%) | 47 (7%) | 55 (8%) | 22 (10%) |
| -6 | 38 (3%) | 22 (3%) | 22 (3%) | 11 (5%) |
| -5 | 60 (5%) | 22 (3%) | 27 (4%) | 9 (4%) |
| -4 | 59 (5%) | 14 (2%) | 29 (4%) | 9 (4%) |
| -3 | 71 (5%) | 41 (6%) | 41 (6%) | 14 (6%) |
| -2 | 85 (7%) | 37 (6%) | 50 (7%) | 17 (8%) |
| -1 | 88 (7%) | 49 (8%) | 49 (7%) | 18 (8%) |
| 0 | 367 (28%) | 179 (28%) | 222 (33%) | 58 (26%) |
| ≥ 1 | 82 (6%) | 40 (6%) | 10 (1%) | 3 (1%) |
| Missing | 191 (15%) | 107 (16%) | 101 (15%) | 34 (16%) |
| **Emergency hospital admission** | | | | |
| Yes | 564 (43%) | 245 (38%) | 447 (66%) | 131 (60%) |
| No | 369 (28%) | 220 (34%) | 223 (33%) | 84 (38%) |
| Missing | 364 (28%) | 184 (28%) | 12 (2%) | 4 (2%) |
| **Month of diagnosis** | | | | |
| ≤3/2020 | 653 (50%) | 333 (51%) | 91 (13%) | 20 (9%) |
| 4/2020 | 495 (38%) | 238 (37%) | 114 (17%) | 23 (11%) |
| 5/2020 | 99 (8%) | 49 (8%) | 29 (4%) | 9 (4%) |
| 6/2020 | 38 (3%) | 23 (4%) | 16 (2%) | 4 (2%) |
| 7/2020 | 12 (1%) | 6 (1%) | 65 (10%) | 37 (17%) |
| 8/2020 | 0 (0%) | 0 (0%) | 77 (11%) | 24 (11%) |
| 9/2020 | 0 (0%) | 0 (0%) | 85 (12%) | 35 (16%) |
| 10/2020 | 0 (0%) | 0 (0%) | 184 (27%) | 59 (27%) |
| 11/2020 | 0 (0%) | 0 (0%) | 21 (3%) | 8 (4%) |
| **Any neurologic comorbidity** | | | | |
| Yes | 291 (22%) | 145 (22%) | 133 (20%) | 32 (15%) |
| No | 774 (60%) | 393 (61%) | 534 (78%) | 182 (83%) |
| Missing | 232 (18%) | 111 (17%) | 15 (2%) | 5 (2%) |
| **Hemiplegia** | | | | |
| Yes | 31 (2%) | 16 (2%) | 8 (1%) | 3 (1%) |
| No | 1235 (95%) | 614 (95%) | 655 (96%) | 211 (96%) |
| Missing | 31 (2%) | 19 (3%) | 19 (3%) | 5 (2%) |
| **Dementia** | | | | |
| Yes | 112 (9%) | 65 (10%) | 53 (8%) | 10 (5%) |
| No | 1153 (89%) | 568 (88%) | 613 (90%) | 204 (93%) |
| Missing | 32 (2%) | 16 (2%) | 16 (2%) | 5 (2%) |
| **Cerebrovascular disease, stroke, TIA** | | | | |
| Yes | 128 (10%) | 63 (10%) | 60 (9%) | 16 (7%) |
| No | 1139 (88%) | 567 (87%) | 604 (89%) | 198 (90%) |
| Missing | 30 (2%) | 19 (3%) | 18 (3%) | 5 (2%) |
| **Movement disorder (e.g. Parkinson's disease, Dystonia, Ataxia, Tremor)** | | | | |
| Yes | 28 (2%) | 12 (2%) | 0 (0%) | 0 (0%) |
| No | 1014 (78%) | 511 (79%) | 0 (0%) | 0 (0%) |
| Missing | 255 (20%) | 126 (19%) | 682 (100%) | 219 (100%) |
| **Other neurological autoimmune diseases** | | | | |
| Yes | 5 (0%) | 3 (0%) | 1 (0%) | 1 (0%) |
| No | 1220 (94%) | 614 (95%) | 662 (97%) | 213 (97%) |
| Missing | 72 (6%) | 32 (5%) | 19 (3%) | 5 (2%) |
| **Other prior neurological diagnosis** | | | | |
| Yes | 72 (6%) | 35 (5%) | 31 (5%) | 9 (4%) |
| No | 970 (75%) | 485 (75%) | 632 (93%) | 205 (94%) |
| Missing | 255 (20%) | 129 (20%) | 19 (3%) | 5 (2%) |
| **Myocardial infarction** | | | | |
| Yes | 80 (6%) | 43 (7%) | 36 (5%) | 7 (3%) |
| No | 1169 (90%) | 583 (90%) | 627 (92%) | 207 (95%) |
| Missing | 48 (4%) | 23 (4%) | 19 (3%) | 5 (2%) |
| **Aortic stenosis** | | | | |
| Yes | 43 (3%) | 18 (3%) | 14 (2%) | 4 (2%) |
| No | 1195 (92%) | 600 (92%) | 648 (95%) | 210 (96%) |
| Missing | 59 (5%) | 31 (5%) | 20 (3%) | 5 (2%) |
| **AV block** | | | | |
| Yes | 36 (3%) | 16 (2%) | 11 (2%) | 2 (1%) |
| No | 1201 (93%) | 603 (93%) | 652 (96%) | 212 (97%) |
| Missing | 60 (5%) | 30 (5%) | 19 (3%) | 5 (2%) |
| **Carotid arterial disease** | | | | |
| Yes | 37 (3%) | 18 (3%) | 12 (2%) | 3 (1%) |
| No | 1184 (91%) | 594 (92%) | 648 (95%) | 211 (96%) |
| Missing | 76 (6%) | 37 (6%) | 22 (3%) | 5 (2%) |
| **Chronic heart failure** | | | | |
| Yes | 132 (10%) | 75 (12%) | 40 (6%) | 14 (6%) |
| No | 1116 (86%) | 550 (85%) | 623 (91%) | 201 (92%) |
| Missing | 49 (4%) | 24 (4%) | 19 (3%) | 4 (2%) |
| **Peripheral vascular disease** | | | | |
| Yes | 59 (5%) | 29 (4%) | 22 (3%) | 7 (3%) |
| No | 1182 (91%) | 589 (91%) | 640 (94%) | 207 (95%) |
| Missing | 56 (4%) | 31 (5%) | 20 (3%) | 5 (2%) |
| **Hypertension** | | | | |
| Yes | 640 (49%) | 310 (48%) | 306 (45%) | 88 (40%) |
| No | 638 (49%) | 331 (51%) | 365 (54%) | 128 (58%) |
| Missing | 19 (1%) | 8 (1%) | 11 (2%) | 3 (1%) |
| **Atrial fibrillation** | | | | |
| Yes | 179 (14%) | 97 (15%) | 78 (11%) | 17 (8%) |
| No | 1083 (84%) | 530 (82%) | 585 (86%) | 198 (90%) |
| Missing | 35 (3%) | 22 (3%) | 19 (3%) | 4 (2%) |
| **Coronary artery disease** | | | | |
| Yes | 190 (15%) | 93 (14%) | 87 (13%) | 19 (9%) |
| No | 1051 (81%) | 526 (81%) | 575 (84%) | 195 (89%) |
| Missing | 56 (4%) | 30 (5%) | 20 (3%) | 5 (2%) |
| **COPD** | | | | |
| Yes | 73 (6%) | 36 (6%) | 41 (6%) | 13 (6%) |
| No | 1198 (92%) | 599 (92%) | 622 (91%) | 200 (91%) |
| Missing | 26 (2%) | 14 (2%) | 19 (3%) | 6 (3%) |
| **Asthma** | | | | |
| Yes | 60 (5%) | 29 (4%) | 47 (7%) | 19 (9%) |
| No | 1209 (93%) | 606 (93%) | 616 (90%) | 194 (89%) |
| Missing | 28 (2%) | 14 (2%) | 19 (3%) | 6 (3%) |
| **Other chronic pulmonary diseases** | | | | |
| Yes | 55 (4%) | 35 (5%) | 25 (4%) | 9 (4%) |
| No | 1194 (92%) | 587 (90%) | 638 (94%) | 204 (93%) |
| Missing | 48 (4%) | 27 (4%) | 19 (3%) | 6 (3%) |
| **Leukemia** | | | | |
| Yes | 23 (2%) | 10 (2%) | 5 (1%) | 1 (0%) |
| No | 1241 (96%) | 625 (96%) | 657 (96%) | 213 (97%) |
| Missing | 33 (3%) | 14 (2%) | 20 (3%) | 5 (2%) |
| **Lymphoma** | | | | |
| Yes | 39 (3%) | 17 (3%) | 9 (1%) | 1 (0%) |
| No | 1226 (95%) | 618 (95%) | 653 (96%) | 213 (97%) |
| Missing | 32 (2%) | 14 (2%) | 20 (3%) | 5 (2%) |
| **Solid tumor** | | | | |
| Yes | 116 (9%) | 60 (9%) | 43 (6%) | 13 (6%) |
| No | 1151 (89%) | 572 (88%) | 618 (91%) | 200 (91%) |
| Missing | 30 (2%) | 17 (3%) | 21 (3%) | 6 (3%) |
| **Solid tumor, metastasized** | | | | |
| Yes | 57 (4%) | 21 (3%) | 17 (2%) | 4 (2%) |
| No | 1205 (93%) | 611 (94%) | 646 (95%) | 210 (96%) |
| Missing | 35 (3%) | 17 (3%) | 19 (3%) | 5 (2%) |
| **Stem cell transplantation** | | | | |
| Yes | 10 (1%) | 7 (1%) | 0 (0%) | 0 (0%) |
| No | 1253 (97%) | 629 (97%) | 663 (97%) | 214 (98%) |
| Missing | 34 (3%) | 13 (2%) | 19 (3%) | 5 (2%) |
| **Connective tissue disease** | | | | |
| Yes | 15 (1%) | 4 (1%) | 5 (1%) | 1 (0%) |
| No | 1250 (96%) | 630 (97%) | 659 (97%) | 213 (97%) |
| Missing | 32 (2%) | 15 (2%) | 18 (3%) | 5 (2%) |
| **Peptic ulcer disease** | | | | |

| Predictor | Deriv. | Valid. | Test, f. | Test, l. |
|---|---|---|---|---|
| Yes | 39 (3%) | 17 (3%) | 5 (1%) | 1 (0%) |
| No | 1226 (95%) | 616 (95%) | 657 (96%) | 213 (97%) |
| Missing | 32 (2%) | 16 (2%) | 20 (3%) | 5 (2%) |
| **Chronic liver disease** | | | | |
| Yes | 29 (2%) | 13 (2%) | 8 (1%) | 3 (1%) |
| No | 1236 (95%) | 622 (96%) | 653 (96%) | 211 (96%) |
| Missing | 32 (2%) | 14 (2%) | 21 (3%) | 5 (2%) |
| **Liver cirrhosis** | | | | |
| Yes | 15 (1%) | 5 (1%) | 6 (1%) | 1 (0%) |
| No | 1252 (97%) | 629 (97%) | 656 (96%) | 213 (97%) |
| Missing | 30 (2%) | 15 (2%) | 20 (3%) | 5 (2%) |
| **Diabetes, no end-organ damage** | | | | |
| Yes | 164 (13%) | 81 (12%) | 101 (15%) | 39 (18%) |
| No | 1106 (85%) | 554 (85%) | 564 (83%) | 177 (81%) |
| Missing | 27 (2%) | 14 (2%) | 17 (2%) | 3 (1%) |
| **Diabetes, with end-organ damage** | | | | |
| Yes | 90 (7%) | 47 (7%) | 35 (5%) | 4 (2%) |
| No | 1177 (91%) | 587 (90%) | 626 (92%) | 210 (96%) |
| Missing | 30 (2%) | 15 (2%) | 21 (3%) | 5 (2%) |
| **Acute kidney injury** | | | | |
| Yes | 59 (5%) | 33 (5%) | 26 (4%) | 5 (2%) |
| No | 888 (68%) | 445 (69%) | 650 (95%) | 214 (98%) |
| Missing | 350 (27%) | 171 (26%) | 6 (1%) | 0 (0%) |
| **Chronic kidney disease** | | | | |
| Yes | 206 (16%) | 97 (15%) | 77 (11%) | 17 (8%) |
| No | 1060 (82%) | 537 (83%) | 587 (86%) | 197 (90%) |
| Missing | 31 (2%) | 15 (2%) | 18 (3%) | 5 (2%) |
| **On dialysis** | | | | |
| Yes | 26 (2%) | 18 (3%) | 14 (2%) | 6 (3%) |
| No | 1241 (96%) | 617 (95%) | 648 (95%) | 208 (95%) |
| Missing | 30 (2%) | 14 (2%) | 20 (3%) | 5 (2%) |
| **Organ transplantation** | | | | |
| Yes | 26 (2%) | 18 (3%) | 10 (1%) | 4 (2%) |
| No | 1240 (96%) | 617 (95%) | 653 (96%) | 210 (96%) |
| Missing | 31 (2%) | 14 (2%) | 19 (3%) | 5 (2%) |
| **Rheumatic disease** | | | | |
| Yes | 46 (4%) | 22 (3%) | 23 (3%) | 5 (2%) |
| No | 1187 (92%) | 598 (92%) | 641 (94%) | 209 (95%) |
| Missing | 64 (5%) | 29 (4%) | 18 (3%) | 5 (2%) |
| **Stage of heart failure (NYHA) at diagnosis** | | | | |
| None | 775 (60%) | 376 (58%) | 501 (73%) | 175 (80%) |
| NYHA I | 48 (4%) | 22 (3%) | 9 (1%) | 3 (1%) |
| NYHA II | 35 (3%) | 28 (4%) | 18 (3%) | 8 (4%) |
| NYHA III | 34 (3%) | 17 (3%) | 24 (4%) | 7 (3%) |
| NYHA IV | 10 (1%) | 5 (1%) | 5 (1%) | 2 (1%) |
| Missing | 395 (30%) | 201 (31%) | 125 (18%) | 24 (11%) |
| **Asymptomatic (sympt.)** | | | | |
| Not quoted | 892 (69%) | 444 (68%) | 503 (74%) | 167 (76%) |
| Quoted | 147 (11%) | 73 (11%) | 77 (11%) | 29 (13%) |
| Missing | 258 (20%) | 132 (20%) | 102 (15%) | 23 (11%) |
| **Runny nose (sympt.)** | | | | |
| Not quoted | 955 (74%) | 481 (74%) | 514 (75%) | 177 (81%) |
| Quoted | 34 (3%) | 21 (3%) | 30 (4%) | 5 (2%) |
| Missing | 308 (24%) | 147 (23%) | 138 (20%) | 37 (17%) |
| **Sore throat (sympt.)** | | | | |
| Not quoted | 935 (72%) | 471 (73%) | 490 (72%) | 170 (78%) |
| Quoted | 59 (5%) | 34 (5%) | 58 (9%) | 14 (6%) |
| Missing | 303 (23%) | 144 (22%) | 134 (20%) | 35 (16%) |
| **Dry cough (sympt.)** | | | | |
| Not quoted | 658 (51%) | 331 (51%) | 353 (52%) | 116 (53%) |
| Quoted | 394 (30%) | 190 (29%) | 260 (38%) | 90 (41%) |
| Missing | 245 (19%) | 128 (20%) | 69 (10%) | 13 (6%) |
| **Productive cough (sympt.)** | | | | |
| Not quoted | 950 (73%) | 471 (73%) | 508 (74%) | 174 (79%) |
| Quoted | 68 (5%) | 34 (5%) | 60 (9%) | 19 (9%) |
| Missing | 279 (22%) | 144 (22%) | 114 (17%) | 26 (12%) |
| **Wheezing (sympt.)** | | | | |
| Not quoted | 993 (77%) | 498 (77%) | 530 (78%) | 178 (81%) |
| Quoted | 16 (1%) | 3 (0%) | 10 (1%) | 4 (2%) |
| Missing | 288 (22%) | 148 (23%) | 142 (21%) | 37 (17%) |
| **Dyspnoe (sympt.)** | | | | |
| Not quoted | 720 (56%) | 359 (55%) | 377 (55%) | 130 (59%) |
| Quoted | 287 (22%) | 143 (22%) | 238 (35%) | 75 (34%) |

| Predictor | Deriv. | Valid. | Test, f. | Test, l. |
|---|---|---|---|---|
| Missing | 290 (22%) | 147 (23%) | 67 (10%) | 14 (6%) |
| **Palpitations (sympt.)** | | | | |
| Not quoted | 989 (76%) | 494 (76%) | 534 (78%) | 176 (80%) |
| Quoted | 15 (1%) | 10 (2%) | 4 (1%) | 2 (1%) |
| Missing | 293 (23%) | 145 (22%) | 144 (21%) | 41 (19%) |
| **Diarrhea (sympt.)** | | | | |
| Not quoted | 909 (70%) | 461 (71%) | 502 (74%) | 170 (78%) |
| Quoted | 110 (8%) | 49 (8%) | 69 (10%) | 23 (11%) |
| Missing | 278 (21%) | 139 (21%) | 111 (16%) | 26 (12%) |
| **Nausea/emesis (sympt.)** | | | | |
| Not quoted | 915 (71%) | 458 (71%) | 501 (73%) | 167 (76%) |
| Quoted | 98 (8%) | 53 (8%) | 61 (9%) | 18 (8%) |
| Missing | 284 (22%) | 138 (21%) | 120 (18%) | 34 (16%) |
| **Muscle aches (sympt.)** | | | | |
| Not quoted | 883 (68%) | 445 (69%) | 470 (69%) | 151 (69%) |
| Quoted | 124 (10%) | 58 (9%) | 77 (11%) | 31 (14%) |
| Missing | 290 (22%) | 146 (22%) | 135 (20%) | 37 (17%) |
| **Muscle weakness (sympt.)** | | | | |
| Not quoted | 895 (69%) | 462 (71%) | 454 (67%) | 147 (67%) |
| Quoted | 109 (8%) | 41 (6%) | 90 (13%) | 35 (16%) |
| Missing | 293 (23%) | 146 (22%) | 138 (20%) | 37 (17%) |
| **Fever (sympt.)** | | | | |
| Not quoted | 580 (45%) | 287 (44%) | 300 (44%) | 101 (46%) |
| Quoted | 461 (36%) | 237 (37%) | 326 (48%) | 103 (47%) |
| Missing | 256 (20%) | 125 (19%) | 56 (8%) | 15 (7%) |
| **Delirium (sympt.)** | | | | |
| Not quoted | 983 (76%) | 493 (76%) | 547 (80%) | 193 (88%) |
| Quoted | 23 (2%) | 10 (2%) | 9 (1%) | 3 (1%) |
| Missing | 291 (22%) | 146 (22%) | 126 (18%) | 23 (11%) |
| **Excessive tiredness (sympt.)** | | | | |
| Not quoted | 808 (62%) | 414 (64%) | 450 (66%) | 144 (66%) |
| Quoted | 210 (16%) | 94 (14%) | 120 (18%) | 50 (23%) |
| Missing | 279 (22%) | 141 (22%) | 112 (16%) | 25 (11%) |
| **Headache (sympt.)** | | | | |
| Not quoted | 890 (69%) | 458 (71%) | 462 (68%) | 157 (72%) |
| Quoted | 109 (8%) | 48 (7%) | 84 (12%) | 25 (11%) |
| Missing | 298 (23%) | 143 (22%) | 136 (20%) | 37 (17%) |
| **Meningism (sympt.)** | | | | |
| Not quoted | 999 (77%) | 501 (77%) | 538 (79%) | 177 (81%) |
| Quoted | 5 (0%) | 2 (0%) | 0 (0%) | 0 (0%) |
| Missing | 293 (23%) | 146 (22%) | 144 (21%) | 42 (19%) |
| **Smelling disorder (sympt.)** | | | | |
| Not quoted | 941 (73%) | 486 (75%) | 503 (74%) | 172 (79%) |
| Quoted | 43 (3%) | 12 (2%) | 26 (4%) | 6 (3%) |
| Missing | 313 (24%) | 151 (23%) | 153 (22%) | 41 (19%) |
| **Tasting disorder (sympt.)** | | | | |
| Not quoted | 926 (71%) | 478 (74%) | 480 (70%) | 164 (75%) |
| Quoted | 58 (4%) | 20 (3%) | 58 (9%) | 14 (6%) |
| Missing | 313 (24%) | 151 (23%) | 144 (21%) | 41 (19%) |
| **Other neurological symptoms** | | | | |
| Not quoted | 959 (74%) | 480 (74%) | 523 (77%) | 173 (79%) |
| Quoted | 56 (4%) | 21 (3%) | 14 (2%) | 6 (3%) |
| Missing | 282 (22%) | 148 (23%) | 145 (21%) | 40 (18%) |
| **Red eye (sympt.)** | | | | |
| Not quoted | 987 (76%) | 498 (77%) | 533 (78%) | 178 (81%) |
| Quoted | 9 (1%) | 6 (1%) | 0 (0%) | 0 (0%) |
| Missing | 301 (23%) | 145 (22%) | 149 (22%) | 41 (19%) |
| **Pulse** | | | | |
| < 60 bpm | 39 (3%) | 16 (2%) | 20 (3%) | 7 (3%) |
| 60 - 89 bpm | 717 (55%) | 359 (55%) | 372 (55%) | 124 (57%) |
| 90 - 119 bpm | 366 (28%) | 180 (28%) | 205 (30%) | 67 (31%) |
| > 119 bpm | 50 (4%) | 26 (4%) | 33 (5%) | 10 (5%) |
| Missing | 125 (10%) | 68 (10%) | 52 (8%) | 11 (5%) |
| **SBP** | | | | |
| < 80 mmHg | 5 (0%) | 3 (0%) | 4 (1%) | 1 (0%) |
| 80 - 99 mmHg | 41 (3%) | 32 (5%) | 20 (3%) | 6 (3%) |
| 100 - 119 mmHg | 282 (22%) | 142 (22%) | 124 (18%) | 37 (17%) |
| 120 - 139 mmHg | 471 (36%) | 223 (34%) | 255 (37%) | 89 (41%) |

| Predictor | Deriv. | Valid. | Test, f. | Test, l. |
|---|---|---|---|---|
| 140 - 179 mmHg | 352 (27%) | 168 (26%) | 210 (31%) | 72 (33%) |
| > 179 mmHg | 33 (3%) | 17 (3%) | 20 (3%) | 4 (2%) |
| Missing | 113 (9%) | 64 (10%) | 49 (7%) | 10 (5%) |
| **DBP** | | | | |
| < 40 mmHg | 3 (0%) | 3 (0%) | 0 (0%) | 0 (0%) |
| 40 - 59 mmHg | 118 (9%) | 60 (9%) | 36 (5%) | 9 (4%) |
| 60 - 89 mmHg | 861 (66%) | 441 (68%) | 480 (70%) | 162 (74%) |
| 90 - 109 mmHg | 188 (14%) | 71 (11%) | 103 (15%) | 32 (15%) |
| > 109 mmHg | 11 (1%) | 7 (1%) | 14 (2%) | 6 (3%) |
| Missing | 116 (9%) | 67 (10%) | 49 (7%) | 10 (5%) |
| **SO2** | | | | |
| < 60% | 10 (1%) | 1 (0%) | 2 (0%) | 1 (0%) |
| 60 - 69% | 5 (0%) | 2 (0%) | 1 (0%) | 0 (0%) |
| 70 - 79% | 14 (1%) | 11 (2%) | 2 (0%) | 1 (0%) |
| 80 - 89% | 124 (10%) | 72 (11%) | 58 (9%) | 16 (7%) |
| 90 - 95% | 487 (38%) | 222 (34%) | 238 (35%) | 67 (31%) |
| 96 - 100% | 505 (39%) | 257 (40%) | 317 (46%) | 118 (54%) |
| Missing | 152 (12%) | 84 (13%) | 64 (9%) | 16 (7%) |
| **Body temperature** | | | | |
| < 35.1 °C | 3 (0%) | 5 (1%) | 0 (0%) | 0 (0%) |
| 35.1 - 37.3 °C | 501 (39%) | 233 (36%) | 282 (41%) | 96 (44%) |
| 37.3 - 37.9 °C | 232 (18%) | 138 (21%) | 139 (20%) | 54 (25%) |
| 38 - 38.9 °C | 310 (24%) | 141 (22%) | 147 (22%) | 36 (16%) |
| 39 - 39.9 °C | 106 (8%) | 63 (10%) | 54 (8%) | 17 (8%) |
| > 39.9 °C | 10 (1%) | 15 (2%) | 6 (1%) | 1 (0%) |
| Missing | 135 (10%) | 54 (8%) | 54 (8%) | 15 (7%) |
| **Respiratory rate** | | | | |
| < 16 | 199 (15%) | 96 (15%) | 110 (16%) | 29 (13%) |
| 16-21 | 411 (32%) | 214 (33%) | 245 (36%) | 85 (39%) |
| 22-29 | 211 (16%) | 91 (14%) | 86 (13%) | 18 (8%) |
| > 29 | 45 (3%) | 29 (4%) | 41 (6%) | 11 (5%) |
| Missing | 431 (33%) | 219 (34%) | 200 (29%) | 76 (35%) |
| **BMI** | | | | |
| < 18.5 kg/m² | 23 (2%) | 17 (3%) | 13 (2%) | 3 (1%) |
| 18.5 - 24.9 kg/m² | 301 (23%) | 152 (23%) | 118 (17%) | 50 (23%) |
| 25 - 29.9 kg/m² | 288 (22%) | 135 (21%) | 159 (23%) | 55 (25%) |
| 30 - 34,9 kg/m² | 121 (9%) | 70 (11%) | 96 (14%) | 42 (19%) |
| > 34,9 kg/m² | 55 (4%) | 23 (4%) | 43 (6%) | 15 (7%) |
| Missing | 509 (39%) | 252 (39%) | 253 (37%) | 54 (25%) |
| **ACE-inhibitors** | | | | |
| Yes | 238 (18%) | 122 (19%) | 147 (22%) | 39 (18%) |
| No | 1022 (79%) | 510 (79%) | 507 (74%) | 169 (77%) |
| Missing | 37 (3%) | 17 (3%) | 28 (4%) | 11 (5%) |
| **AT1-RB** | | | | |
| Yes | 217 (17%) | 108 (17%) | 98 (14%) | 29 (13%) |
| No | 1025 (79%) | 524 (81%) | 544 (80%) | 178 (81%) |
| Missing | 55 (4%) | 17 (3%) | 40 (6%) | 12 (5%) |
| **Statines** | | | | |
| Yes | 253 (20%) | 138 (21%) | 136 (20%) | 34 (16%) |
| No | 770 (59%) | 378 (58%) | 512 (75%) | 174 (79%) |
| Missing | 274 (21%) | 133 (20%) | 34 (5%) | 11 (5%) |
| **Ibuprofen** | | | | |
| Yes | 47 (4%) | 28 (4%) | 29 (4%) | 5 (2%) |
| No | 1129 (87%) | 566 (87%) | 586 (86%) | 194 (89%) |
| Missing | 121 (9%) | 55 (8%) | 67 (10%) | 20 (9%) |
| **Immunosuppressants** | | | | |
| Yes | 131 (10%) | 71 (11%) | 51 (7%) | 19 (9%) |
| No | 1014 (78%) | 515 (79%) | 571 (84%) | 194 (89%) |
| Missing | 152 (12%) | 63 (10%) | 60 (9%) | 6 (3%) |
| **Complementary/integrative medicine** | | | | |
| Yes | 18 (1%) | 4 (1%) | 12 (2%) | 0 (0%) |
| No | 861 (66%) | 438 (67%) | 593 (87%) | 213 (97%) |

| Predictor | Deriv. | Valid. | Test, f. | Test, l. |
|---|---|---|---|---|
| Missing | 418 (32%) | 207 (32%) | 77 (11%) | 6 (3%) |
| **Smoking status** | | | | |
| 1-10 pack years | 22 (2%) | 5 (1%) | 8 (1%) | 3 (1%) |
| 11-20 pack years | 14 (1%) | 8 (1%) | 6 (1%) | 2 (1%) |
| >21 pack years | 34 (3%) | 13 (2%) | 9 (1%) | 2 (1%) |
| Nonsmoker | 468 (36%) | 255 (39%) | 222 (33%) | 84 (38%) |
| Former smoker | 103 (8%) | 57 (9%) | 50 (7%) | 13 (6%) |
| Smoker, unknown pack years | 21 (2%) | 14 (2%) | 24 (4%) | 9 (4%) |
| Missing | 635 (49%) | 297 (46%) | 363 (53%) | 106 (48%) |
| **IgG-titer positive** | | | | |
| Yes | 675 (52%) | 349 (54%) | 645 (95%) | 192 (88%) |
| No | 608 (47%) | 293 (45%) | 0 (0%) | 0 (0%) |
| 51 | 0 (0%) | 0 (0%) | 15 (2%) | 12 (5%) |
| Missing | 14 (1%) | 7 (1%) | 22 (3%) | 15 (7%) |
| **AST** | | | | |
| < LLN | 2 (0%) | 5 (1%) | 9 (1%) | 1 (0%) |
| Normal (LLN - ULN) | 714 (55%) | 336 (52%) | 367 (54%) | 139 (63%) |
| > ULN ≤ 2x ULN | 283 (22%) | 160 (25%) | 133 (20%) | 60 (27%) |
| > 2x ULN ≤ 5x ULN | 73 (6%) | 46 (7%) | 33 (5%) | 12 (5%) |
| > 5x ULN ≤ 10x ULN | 8 (1%) | 3 (0%) | 3 (0%) | 0 (0%) |
| Missing | 217 (17%) | 99 (15%) | 137 (20%) | 7 (3%) |
| **ALT** | | | | |
| < LLN | 7 (1%) | 8 (1%) | 9 (1%) | 2 (1%) |
| Normal (LLN - ULN) | 859 (66%) | 416 (64%) | 470 (69%) | 162 (74%) |
| > ULN ≤ 2x ULN | 220 (17%) | 108 (17%) | 100 (15%) | 38 (17%) |
| > 2x ULN ≤ 5x ULN | 42 (3%) | 34 (5%) | 31 (5%) | 10 (5%) |
| > 5x ULN ≤ 10x ULN | 8 (1%) | 6 (1%) | 5 (1%) | 1 (0%) |
| Missing | 161 (12%) | 77 (12%) | 67 (10%) | 6 (3%) |
| **Gamma-GT** | | | | |
| Normal (LLN - ULN) | 678 (52%) | 349 (54%) | 387 (57%) | 136 (62%) |
| > ULN ≤ 2x ULN | 222 (17%) | 126 (19%) | 108 (16%) | 41 (19%) |
| > 2x ULN ≤ 5x ULN | 106 (8%) | 56 (9%) | 47 (7%) | 25 (11%) |
| > 5x ULN ≤ 10x ULN | 34 (3%) | 8 (1%) | 15 (2%) | 3 (1%) |
| > 10x ULN | 0 (0%) | 0 (0%) | 9 (1%) | 1 (0%) |
| Missing | 257 (20%) | 110 (17%) | 116 (17%) | 13 (6%) |
| **Bilirubine** | | | | |
| < LLN | 7 (1%) | 7 (1%) | 14 (2%) | 2 (1%) |
| Normal (LLN - ULN) | 928 (72%) | 470 (72%) | 516 (76%) | 196 (89%) |
| > ULN ≤ 2x ULN | 50 (4%) | 31 (5%) | 41 (6%) | 12 (5%) |
| > 2x ULN ≤ 5x ULN | 9 (1%) | 4 (1%) | 5 (1%) | 0 (0%) |
| Missing | 303 (23%) | 137 (21%) | 106 (16%) | 9 (4%) |
| **Lipase** | | | | |
| < LLN | 5 (0%) | 2 (0%) | 12 (2%) | 4 (2%) |
| Normal (LLN - ULN) | 463 (36%) | 223 (34%) | 342 (50%) | 152 (69%) |
| > ULN ≤ 2x ULN | 96 (7%) | 51 (8%) | 49 (7%) | 15 (7%) |
| > 2x ULN ≤ 5x ULN | 11 (1%) | 13 (2%) | 8 (1%) | 1 (0%) |
| Missing | 722 (56%) | 360 (55%) | 271 (40%) | 47 (21%) |
| **Troponine T** | | | | |
| < LLN | 7 (1%) | 8 (1%) | 14 (2%) | 8 (4%) |

| Predictor | Deriv. | Valid. | Test, f. | Test, l. |
|---|---|---|---|---|
| Normal (LLN - ULN) | 431 (33%) | 221 (34%) | 250 (37%) | 126 (58%) |
| > ULN ≤ 2x ULN | 84 (6%) | 43 (7%) | 49 (7%) | 24 (11%) |
| > 2x ULN ≤ 5x ULN | 57 (4%) | 28 (4%) | 43 (6%) | 16 (7%) |
| > 5x ULN ≤ 10x ULN | 18 (1%) | 12 (2%) | 12 (2%) | 1 (0%) |
| > 10x ULN ≤ 20x ULN | 10 (1%) | 7 (1%) | 4 (1%) | 2 (1%) |
| > 20x ULN | 3 (0%) | 4 (1%) | 6 (1%) | 1 (0%) |
| Missing | 687 (53%) | 326 (50%) | 304 (45%) | 41 (19%) |
| **Creatinine** | | | | |
| < LLN | 15 (1%) | 13 (2%) | 32 (5%) | 6 (3%) |
| Normal (LLN - ULN) | 918 (71%) | 443 (68%) | 477 (70%) | 174 (79%) |
| > ULN ≤ 2x ULN | 257 (20%) | 140 (22%) | 122 (18%) | 27 (12%) |
| > 2x ULN ≤ 5x ULN | 69 (5%) | 28 (4%) | 26 (4%) | 4 (2%) |
| > 5x ULN ≤ 20x ULN | 12 (1%) | 10 (2%) | 12 (2%) | 7 (3%) |
| Missing | 26 (2%) | 15 (2%) | 13 (2%) | 1 (0%) |
| **Leukocytes** | | | | |
| < 1,000 /µL | 6 (0%) | 5 (1%) | 3 (0%) | 1 (0%) |
| 1,000 - 3999 /µL | 234 (18%) | 116 (18%) | 105 (15%) | 35 (16%) |
| 4,000 - 7,999 /µL | 742 (57%) | 365 (56%) | 386 (57%) | 132 (60%) |
| 8,000 - 11,999 /µL | 224 (17%) | 113 (17%) | 124 (18%) | 38 (17%) |
| 12,000 - 15,999 /µL | 43 (3%) | 27 (4%) | 34 (5%) | 8 (4%) |
| 16,000 - 19,999 /µL | 22 (2%) | 10 (2%) | 10 (1%) | 0 (0%) |
| ≥ 20,000 /µL | 15 (1%) | 10 (2%) | 14 (2%) | 4 (2%) |
| Missing | 11 (1%) | 3 (0%) | 6 (1%) | 1 (0%) |
| **Platelets** | | | | |
| < 10,000 /µL | 7 (1%) | 2 (0%) | 3 (0%) | 1 (0%) |
| 10,000 - 49,999 /µL | 22 (2%) | 9 (1%) | 7 (1%) | 4 (2%) |
| 50,000 - 119,999 /µL | 121 (9%) | 64 (10%) | 59 (9%) | 18 (8%) |
| 120,000 - 449,999 /µL | 1084 (84%) | 554 (85%) | 579 (85%) | 193 (88%) |
| 450,000 - 799,000 /µL | 29 (2%) | 12 (2%) | 15 (2%) | 1 (0%) |
| Missing | 34 (3%) | 8 (1%) | 19 (3%) | 2 (1%) |
| **Hemoglobin** | | | | |
| < 6 g/dL | 6 (0%) | 3 (0%) | 0 (0%) | 0 (0%) |
| 6 - 7.9 g/dL | 40 (3%) | 26 (4%) | 14 (2%) | 4 (2%) |
| 8 - 9.9 g/dL | 115 (9%) | 48 (7%) | 40 (6%) | 13 (6%) |
| 10 - 11.9 g/dL | 243 (19%) | 109 (17%) | 137 (20%) | 37 (17%) |
| 12 - 14.9 g/dL | 677 (52%) | 364 (56%) | 375 (55%) | 132 (60%) |
| ≥ 15 g/dL | 197 (15%) | 94 (14%) | 107 (16%) | 32 (15%) |
| Missing | 19 (1%) | 5 (1%) | 9 (1%) | 1 (0%) |
| **Areas of consolidation (CT)** | | | | |
| Not quoted | 521 (40%) | 278 (43%) | 428 (63%) | 127 (58%) |
| Quoted | 216 (17%) | 103 (16%) | 115 (17%) | 55 (25%) |
| Missing | 560 (43%) | 268 (41%) | 139 (20%) | 37 (17%) |
| **Crazy paving pattern (CT)** | | | | |
| Not quoted | 656 (51%) | 332 (51%) | 513 (75%) | 165 (75%) |
| Quoted | 37 (3%) | 29 (4%) | 30 (4%) | 17 (8%) |
| Missing | 604 (47%) | 288 (44%) | 139 (20%) | 37 (17%) |
| **Ground glass opacities (CT)** | | | | |
| Not quoted | 376 (29%) | 208 (32%) | 329 (48%) | 70 (32%) |
| Quoted | 361 (28%) | 173 (27%) | 214 (31%) | 112 (51%) |
| Missing | 560 (43%) | 268 (41%) | 139 (20%) | 37 (17%) |
| **Interlobular septal thickening (CT)** | | | | |
| Not quoted | 664 (51%) | 348 (54%) | 529 (78%) | 174 (79%) |
| Quoted | 29 (2%) | 13 (2%) | 14 (2%) | 8 (4%) |
| Missing | 604 (47%) | 288 (44%) | 139 (20%) | 37 (17%) |
| **Nodulary lesions (CT)** | | | | |
| Not quoted | 707 (55%) | 364 (56%) | 521 (76%) | 164 (75%) |
| Quoted | 30 (2%) | 17 (3%) | 22 (3%) | 18 (8%) |
| Missing | 560 (43%) | 268 (41%) | 139 (20%) | 37 (17%) |
| **Pleural effusion (CT)** | | | | |
| Not quoted | 686 (53%) | 354 (55%) | 509 (75%) | 170 (78%) |
| Quoted | 51 (4%) | 27 (4%) | 34 (5%) | 12 (5%) |
| Missing | 560 (43%) | 268 (41%) | 139 (20%) | 37 (17%) |
| **Normal (CT)** | | | | |
| Not quoted | 679 (52%) | 337 (52%) | 487 (71%) | 168 (77%) |
| Quoted | 58 (4%) | 44 (7%) | 56 (8%) | 14 (6%) |
| Missing | 560 (43%) | 268 (41%) | 139 (20%) | 37 (17%) |
| **MDRO colonization** | | | | |
| Yes | 55 (4%) | 19 (3%) | 16 (2%) | 6 (3%) |
| No | 748 (58%) | 395 (61%) | 0 (0%) | 0 (0%) |
| Missing | 494 (38%) | 235 (36%) | 666 (98%) | 213 (97%) |

Abbreviations: ACE, angiotensin-converting-enzyme; AT1-RB, angiotensin II receptor type 1 antagonists; AV, atrioventricular; BMI, body mass index; bpm, beats per minute; COPD, chronic obstructive pulmonary disease; CT, computed tomography; DBP, diastolic blood pressure; LLN, lower limit of normal; MDRO, multidrug-resistant organism; NYHA, New York Heart Association functional classification; SBP, systolic blood pressure; sO2, oxygen saturation (%); sympt., variable describes whether a COVID-19 symptom is present at diagnosis; TIA, transient ischemic attack; ULN, upper limit of normal.

### Supplementary Table 2. Summary of the predictive performances of Random Survival Forest and Cox models in comparison to the five variable score.

| | | | C-index (imp. range) | |
|---|---|---|---|---|
| **Selection** | **Model** | **N pr.** | **Derivation** | **Validation** |
| All pr. | RSF | 105 | 0.78 (0.78-0.78) | 0.78 (0.77-0.78) |
| | Cox ridge | 105 | 0.83 (0.81-0.83) | 0.77 (0.77-0.77) |
| RSF Boruta | RSF | 24 | 0.79 (0.79-0.80) | 0.77 (0.76-0.77) |
| | Cox ridge | 24 | 0.82 (0.81-0.82) | 0.77 (0.77-0.77) |
| RF Boruta | RSF | 5 | 0.74 (0.73-0.75) | 0.73 (0.72-0.74) |
| | Cox ridge | 5 | 0.76 (0.76-0.76) | 0.76 (0.76-0.77) |
| | Binomial ridge | 5 | 0.76 (0.76-0.76) | 0.76 (0.76-0.77) |
| | Score | 5 | 0.76 (0.76-0.76) 95%CI, 0.73-0.79 | 0.76 (0.76-0.77) 95%CI, 0.73-0.80 |
| | Validation on full test set | | | 0.80 (0.80-0.81) 95%CI, 0.76-0.84 |
| | Validation on limited test set | | | 0.81 (0.81-0.81) 95%CI, 0.74-0.87 |

Initial derivation and validation analyses were performed on the respective datasets (n=1297 and 649, respectively) as summarized in Figure 2. As indicated, the final score was additionally independently validated on the full and the limited test sets (n=675 and 218, as described in Figure 2).
Indicated are the median values and the full range for the imputed datasets (in brackets). Harrell's C-indices were calculated using a time to event variable (as summarized in Supplementary Figure 1) in addition to the outcome. 95% confidence intervals (95%CI) were calculated for score predictions using bootstrapping with equal contributions of the imputed datasets.
Abbreviations: AUC, area under the receiver operating characteristic (ROC) curve; C-index, concordance index; imp., imputation; N pr., number of predictors in the model; pr., predictors; RSF, random survival forest.

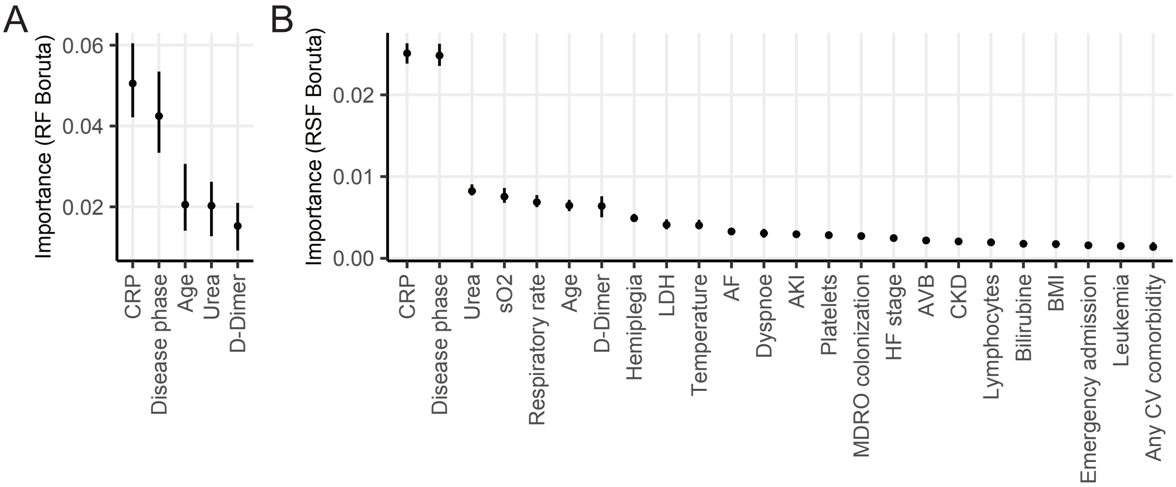


**Supplementary Figure 1. Summary of the selected variables in the RF Boruta (A) and RSF Boruta (B) approaches.** Indicated are the medians and interquartile ranges of variable importance obtained during the respective run of Boruta selection. Abbreviations: sO2, blood oxygen saturation; LDH, lactate dehydrogenase; AF, atrial fibrillation; AKI, acute kidney injury; AVB, atrioventricular block; CKD, chronic kidney disease; BMI, body mass index; CV, cardiovascular.

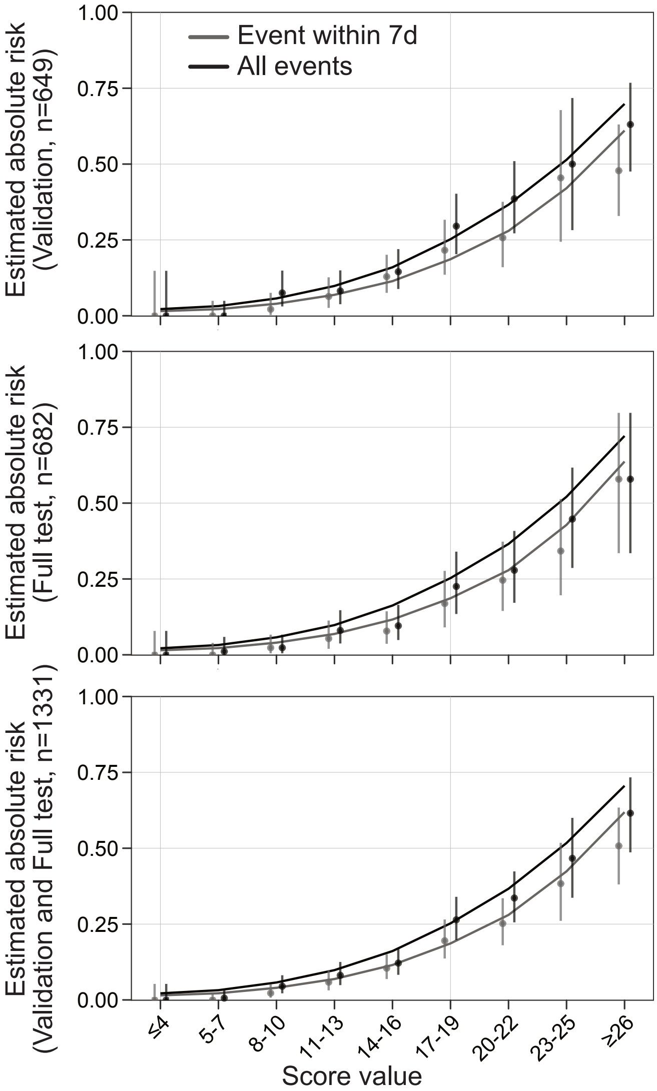


**Supplementary Figure 2. Absolute risks for an event within 7 days of diagnosis or for all events for the indicated validation/test datasets.** Lines represent mean predicted risk by a model calibrated on the derivation dataset. Dots represent observed absolute risk in the respective cohorts. Ranges indicate CI 95%.

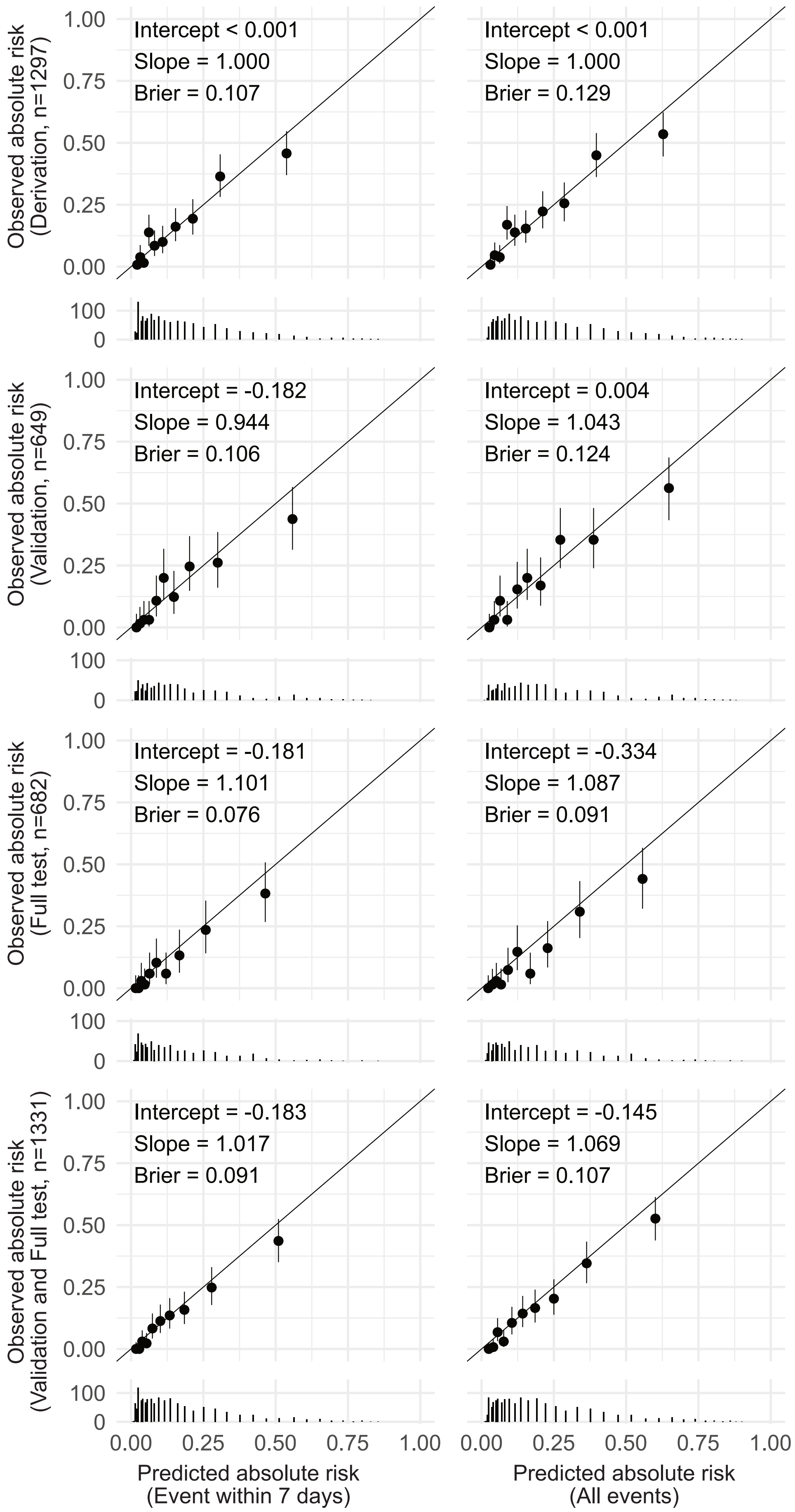


**Supplementary Figure 3. Score calibration results for event risk within 7 days of diagnosis (left panels) or for all events (right panels).** Predictions for the indicated datasets were calculated with models calibrated on the derivation dataset. Solid lines represent optimal calibration. Ranges indicate CI 95%.

STROBE Statement—Checklist of items that should be included in reports of ***cohort studies***

| | Item No | Recommendation | Page No |
|---|---|---|---|
| **Title and abstract** | 1 | (*a*) Indicate the study's design with a commonly used term in the title or the abstract | 2 |
| | | (*b*) Provide in the abstract an informative and balanced summary of what was done and what was found | 2 |
| **Introduction** | | | |
| Background/rationale | 2 | Explain the scientific background and rationale for the investigation being reported | 2 |
| Objectives | 3 | State specific objectives, including any prespecified hypotheses | 2-3 |
| **Methods** | | | |
| Study design | 4 | Present key elements of study design early in the paper | 3 |
| Setting | 5 | Describe the setting, locations, and relevant dates, including periods of recruitment, exposure, follow-up, and data collection | 3 |
| Participants | 6 | (*a*) Give the eligibility criteria, and the sources and methods of selection of participants. Describe methods of follow-up | 3[1] |
| | | (*b*) For matched studies, give matching criteria and number of exposed and unexposed | n.a. |
| Variables | 7 | Clearly define all outcomes, exposures, predictors, potential confounders, and effect modifiers. Give diagnostic criteria, if applicable | 3[1], T.1, S.T.1 |
| Data sources/ measurement | 8* | For each variable of interest, give sources of data and details of methods of assessment (measurement). Describe comparability of assessment methods if there is more than one group | 3[1] |
| Bias | 9 | Describe any efforts to address potential sources of bias | 4 |
| Study size | 10 | Explain how the study size was arrived at | 3 |
| Quantitative variables | 11 | Explain how quantitative variables were handled in the analyses. If applicable, describe which groupings were chosen and why | 3 |
| Statistical methods | 12 | (*a*) Describe all statistical methods, including those used to control for confounding | 3-4 |
| | | (*b*) Describe any methods used to examine subgroups and interactions | n.a. |
| | | (*c*) Explain how missing data were addressed | 4 |
| | | (*d*) If applicable, explain how loss to follow-up was addressed | 4 |
| | | (*e*) Describe any sensitivity analyses | F. 3 |
| **Results** | | | |
| Participants | 13* | (a) Report numbers of individuals at each stage of study—eg numbers potentially eligible, examined for eligibility, confirmed eligible, included in the study, completing follow-up, and analysed | 3, F.2 |
| | | (b) Give reasons for non-participation at each stage | 3, F.2 |
| | | (c) Consider use of a flow diagram | F.2 |
| Descriptive data | 14* | (a) Give characteristics of study participants (eg demographic, clinical, social) and information on exposures and potential confounders | T.1, S.T.1 |
| | | (b) Indicate number of participants with missing data for each variable of interest | T.1, S.T.1 |
| | | (c) Summarise follow-up time (eg, average and total amount) | S.T.1 |
| Outcome data | 15* | Report numbers of outcome events or summary measures over time | T.1 |

| | | | |
|---|---|---|---|
| Main results | 16 | (*a*) Give unadjusted estimates and, if applicable, confounder-adjusted estimates and their precision (eg, 95% confidence interval). Make clear which confounders were adjusted for and why they were included<br>(*b*) Report category boundaries when continuous variables were categorized<br>(*c*) If relevant, consider translating estimates of relative risk into absolute risk for a meaningful time period | T.2, T.3, T.5<br>T. 1 S.T.1 S.F.2 |
| Other analyses | 17 | Report other analyses done—eg analyses of subgroups and interactions, and sensitivity analyses | F. 3, S.F.3 |
| **Discussion** | | | |
| Key results | 18 | Summarise key results with reference to study objectives | 6-8 |
| Limitations | 19 | Discuss limitations of the study, taking into account sources of potential bias or imprecision. Discuss both direction and magnitude of any potential bias | 8-9 |
| Interpretation | 20 | Give a cautious overall interpretation of results considering objectives, limitations, multiplicity of analyses, results from similar studies, and other relevant evidence | 6-9 |
| Generalisability | 21 | Discuss the generalisability (external validity) of the study results | 7-9 |
| **Other information** | | | |
| Funding | 22 | Give the source of funding and the role of the funders for the present study and, if applicable, for the original study on which the present article is based | 9 |

*Give information separately for exposed and unexposed groups.

Abbreviations: F., figure; T., table; S., supplementary.
[1]We additionally reference a previous publications from the LEOSS group which reports on the study design in further detail: Jakob et al., First results of the “Lean European Open Survey on SARS-CoV-2-Infected Patients (LEOSS)”. Infection 2020.

**Note:** An Explanation and Elaboration article discusses each checklist item and gives methodological background and published examples of transparent reporting. The STROBE checklist is best used in conjunction with this article (freely available on the Web sites of PLoS Medicine at http://www.plosmedicine.org/, Annals of Internal Medicine at http://www.annals.org/, and Epidemiology at http://www.epidem.com/). Information on the STROBE Initiative is available at http://www.strobe-statement.org.
